# Multiple imputation strategies for missing event times in a multi-state model analysis

**DOI:** 10.1101/2023.06.16.23291499

**Authors:** E. Curnow, R. A. Hughes, K. Birnie, K. Tilling, M. J. Crowther

## Abstract

In clinical studies, multi-state model (MSM) analysis is often used to describe the sequence of events that patients experience, enabling better understanding of disease progression. A complicating factor in many MSM studies is that the exact event times may not be known. Motivated by a real dataset of patients who received stem cell transplants, we considered the setting in which some event times were exactly observed and some were missing. In our setting, there was little information about the time intervals in which the missing event times occurred and missingness depended on the event type, given the analysis model covariates. These additional challenges limited the usefulness of some missing data methods (maximum likelihood, complete case analysis, and inverse probability weighting). We show, for the first time in the MSM context, that multiple imputation (MI) of event times can perform well in this setting. MI is a flexible method that can be used with any complete data analysis model. Through an extensive simulation study, we show that MI by predictive mean matching (PMM), in which sampling is from a set of observed times without reliance on a specific parametric distribution, has little bias when event times are missing at random, conditional on the observed data. Applying PMM separately for each sub-group of patients with a different pathway through the MSM tends to further reduce bias and improve precision. We recommend MI using PMM methods when performing MSM analysis with Markov models and partially observed event times.

## 1. Introduction

In clinical studies, there is often interest in describing the sequence of events that each patient experiences, to enable better understanding of disease progression. Increasingly, multi-state model (MSM) analysis is used for this purpose. MSMs have been used in a wide variety of clinical contexts, such as organ and stem cell transplantation,^1, 2^ studies of dementia^3^ and aging,^4^ and in cancer research.^5^ The advantage of the MSM approach is that the probability of multiple events can be modelled simultaneously. This allows the prediction of clinically-relevant quantities, such as the probability of each event at any given time, and the average number of days spent in each state. This in turn enables more effective communication of risk to patients,^6^ particularly because these quantities can easily be illustrated graphically.

A complicating factor in MSM studies is that the exact time of each event may not be known. In some settings, none of the event times are exactly observed (with the possible exception of time of death). For example, in HIV^7^ or dentistry,^8^ changes in the health of the patient are reported only at intermittent clinic visits. Formally, such events are “interval-censored”: the event time lies in the interval (*L*, *R*], where *L* represents the last known event-free time and *R* represents the first time at which the event is reported. In such settings, maximum likelihood (ML) methods for interval-censored data^1, 3, 9^ - in which the marginal likelihood of the observed data is maximised - are generally used. In other settings, exact event times are observed for some individuals but not others. For example, in a pregnancy study,^10^ gestational age at delivery was recorded for some individuals but missing for others. In this type of setting, a wider choice of methods for missing data is available because some individuals have complete data. As well as ML, available methods include complete case analysis (CCA), inverse probability weighting (IPW), and multiple imputation (MI).^11^ Our motivating example is in this type of setting. We consider a previously analysed dataset of patients who received haematopoietic stem cell (HSC) transplants using cord blood (CB) donated to the UK National Health Service (NHS) Cord Blood Bank (CBB).^12^ There were missing data in the NHS CBB dataset. In particular, the times of onset of acute graft-versus-host disease^13^ (aGvHD, caused by an immune response of donor cells - the “graft” - against the patient’s tissues and organs - the “host”) and relapse (*i.e.* signs and symptoms that the patient’s original blood disease has returned after treatment) were missing for approximately 25% of patients who experienced aGvHD and/or relapse, respectively. Exact times of death or last follow-up were reported for all patients. Note that these missing times can still be considered interval-censored, with finite interval boundaries inferred from clinical criteria (*e.g.* the standard clinical definition of aGvHD^13^ assumes occurrence between day 0 and 100 post-transplant) or the known length of the monitoring period for each patient.

The NHS CBB dataset is an interesting test case because ML, CCA, and IPW have limited use, for the following reasons:

i. Our setting deviates from the assumptions of available ML methods in two ways:

1. Our event times are a mixture of observed and missing (interval-censored) times. ML methods developed so far assume that all times are interval-censored.
2. For our missing times, the associated interval boundaries are wide relative to the observed event times. In a review of ML methods for handling interval-censoring in MSM analysis, Machado *et al.* found that none of the available methods performed well when censoring intervals were wide, relative to the change in hazards.^14^
ii. CCA (in which only patients with observed values for all analysis model variables are included) will give biased estimates in our setting because missingness depends on the analysis model outcome.^15^ Note that CCA estimates would only be unbiased in this setting if the probability that event times were missing did not depend on the type of event nor the event times themselves (after conditioning on the analysis model covariates).
iii. IPW (in which the complete cases are weighted by the inverse of the estimated probability of being complete) is likely to perform poorly because the types of event experienced by each patient is strongly predictive of missingness of the event times, resulting in extreme weights for some individuals.^16^ In addition, like CCA, IPW estimates generally lack precision because the incomplete cases (which contain partial information about the outcome) are discarded.^11^

In contrast to approaches (i)-(iii), above, MI (assuming the missingness mechanism is ignorable and the imputation model is correctly specified^17^) utilises all available data, using observed data for the analysis model variables plus any additional variables that are predictive of the missing event times, from both patients with fully observed event times and those with partially observed event times. In addition, MI can accommodate a mixture of exactly observed and missing times, plus it allows flexibility when choosing the analysis model.

The standard MI procedure consists of three steps:

1. An imputation model is fitted to the observed data and missing values are replaced with draws from its predictive distribution. This is repeated multiple (M) times, to give M completed datasets.
2. The analysis model is fitted to each of the M completed datasets.
3. The M sets of results are combined using Rubin’s rules.^18^

Using the NHS CBB dataset as motivation, Curnow *et al.*^12^ considered MI and ML strategies for handling missing event times in a competing risks analysis. They examined the extent to which interval boundaries, the data distribution, and analysis model should be accounted for in the imputation model. They found that MI by predictive mean matching^19^ (PMM) resulted in least biased estimates, and was robust to model mis-specification. PMM is a variation on the standard MI procedure described above, in which missing values are replaced with observed values from donors with a similar predicted mean. Therefore, in this paper, we also focus on MI, extending the work of Curnow *et al.* to multi-state Markov models. This is the first time that the performance of MI methods for missing event times has been assessed in the MSM context. Note that, in this paper, we generally refer to ‘event’ times rather than ‘transition’ times. This is because it is more realistic to have missing times for a specific event - which may affect several transition times – rather than for a specific transition (*e.g.* a missing times of aGvHD will affect times of transition to and from aGvHD).

In Section 2 we describe the motivating example. In Section 3 we describe MSM methodology in detail. In Section 4 we describe a simulation study comparing different MI strategies and present its results in Section 5. In Section 6 we apply our MI strategies to the motivating study dataset. We conclude with general discussion in Section 7.

## 2. Motivating Study

The NHS CBB dataset contained information about 432 CB transplants. Individual-level data were available about baseline patient, donor, and transplant characteristics (see Supplementary Material Section S4 for further details) as well as about events experienced by each patient during the post-transplant monitoring period. Event types included aGvHD and chronic GvHD (cGvHD, GvHD occurring more than 100 days post-transplant), relapse, and death. The median follow-up time was 3 years (Kaplan-Meier estimate, censoring follow-up time at death) and at least one post-transplant event was reported for each patient. For each type of event, both an indicator of whether the event was experienced and the associated time of onset were reported (censoring at the earliest of the time of a competing event or last follow-up). The time of onset of aGvHD was missing for 57 (24%) of 241 patients who experienced aGvHD, and the time of onset of relapse was missing for 22 (25%) of 89 patients who experienced relapse.

## 3. Multi-state models

Formally, we consider a stochastic process {*Y*(*t*), *t* ∈ *T*} with a finite state space *Z* = {0, …, *N*} and process history up to time *s*, *Hs* = {*Y*(*u*); 0 ≤ *u* ≤ *s*}.^2^ Then *P*(*Y*(*t*) *= b* | *Y*(*s*) = *a*, *Hs*) represents the probability that a patient in state *a* at time *s* moves to state *b* at time *t*, given the process history up to time *s*, where *a*, *b* ∈ *Z*. Analogous to the hazard rate in standard survival analysis, the transition intensity, α*_ab_*(*t*), is defined as the instantaneous probability of moving from state *a* to state *b* at time *t*. The set of transition intensities fully characterises the multi-state process.

MSMs are often represented using diagrams, such as Figure 1 below, which depicts the classic “illness-death” model. In Figure 1, states are represented by rectangles, and possible transitions by arrows. The transition intensity, α*_ab_*(*t*), is shown for each transition. In Figure 1, there is a single initial state (0. Alive), and we assume that all patients are in this state at the time origin, *t* = 0. There is a single “absorbing” state (2. Death), that is, a state from which further transitions cannot occur. There is also an “intermediate” state (1. Illness) between initial and absorbing states. There are two possible pathways through this MSM: either 0-1-2 (patient *i* is alive without illness at *t* = 0, becomes ill at time *t_1i_*, and dies at time *t_2i_*, with 0 < *t_1i_* < *t_2i_*), or 0-2 (patient *j* is alive without illness at *t* = 0, and is still without illness at time of death *t_2j_*, with *t_2j_* > 0). Patients can be right-censored at any time-point along their pathway. More complex MSMs may have mutiple initial, intermediate or absorbing states, and bi-directional arrows, but these types of MSM are out of scope for this paper.

**Figure 1.**
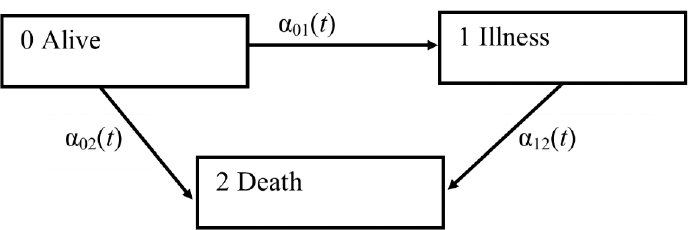
The illness-death multi-state model

The calculation of transition intensities and related probabilities is most straight-forward for MSMs with the Markov property.^20^ This property states that the transition probability depends only on the current state occupied but not the amount of time spent in the current state nor the past history prior to entry into the current state. Hence, in this case, the transition probability can be simplified to *P*(*Y*(*t*) *= b* | *Y*(*s*) = *a*), hereafter denoted by *P_ab_*(*s,t*). For a Markov model, the matrix of transition probabilities, **P**(*s,t*), is calculated from the transition intensities as follows: **P**(*s,t*) = ∏_(_*_s,t_*_]_(**I** + *d**H**(u)*) where ***H****(u)* is the matrix of cumulative transition intensities.^21^ This paper only considers Markov models. We include a test for the Markov property in the simulation and real data analysis.

## 4. Simulation study

We conducted a simulation study to assess the performance of MI methods when applied to a MSM analysis, when some event times were missing. The aim of the simulation study was to quantify the bias and precision of estimates from a MSM analysis in various missing data scenarios. The design of the simulation study is summarised below. Further details are provided in Supplementary Material Section S1.

### 4.1. Data generation

We first generated complete data for the event times and associated states using the method described by Beyersmann *et al.*^22^ applied to a Markov uni-directional three-state model (similar to Figure 1 above). We used 1000 simulations and each simulated dataset contained 500 patients (similar to the size of the real dataset). In our model, transplant was the initial state for all patients (state 0), aGvHD the single intermediate state (state 1), and relapse or death (a composite outcome which is of clinical interest in many HSC transplant studies^23–25^) was the single absorbing state (state 2). Note that here, for simplicity, we generated a single time of relapse/death for each patient (rather than separately generating a time of relapse and a time of death as in the real data).

We assumed that each transition intensity model, α*_ab_*(*t*), had a proportional hazards (PH) structure, defined as follows:

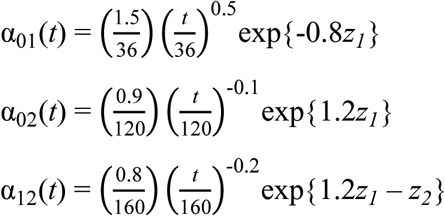

where *t* represents the time in days since transplant, *z_1_*∼ *Bernoulli*(0.2) represents whether a patient is in relapse at time of transplant (assuming patients in relapse at the time of transplant are relapse-free immediately post-transplant), *z_2_* ∼ *Bernoulli*(0.45) represents whether a patient receives a double cord transplant (vs. single cord), and *z_1_*and *z_2_* are independent. The magnitude of the model parameters and choice of covariates were based on the real data. Censoring times were randomly generated between one and five years post-transplant, to represent administrative (non-informative) censoring at study end.

In a subsequent step, missing event times were generated using 12 different missing data mechanisms (MDMs). Firstly, we considered event times missing completely at random (MCAR, *i.e.* missingness was independent of both the observed and missing data) by setting a random 30% of event times to missing, regardless of the event type. Next, we considered 11 different MDMs (see Supplementary Material Table S1). We assumed that event times to acute GvHD and/or relapse/death were either (i) missing at random (MAR), conditional on the observed data (missingness depended on the event type and covariates but not on the missing data itself), or (ii) missing not at random (MNAR, missingness depended on the missing data itself). Although our chosen MI methods assumed data were MAR, MNAR MDMs allowed us to assess the impact on bias and precision when the MAR assumption was violated. Approximately 30% of event times were missing in each MDM, to reflect the percentage of missing times in the real data.

### 4.2. Analysis model, estimands and performance measures

Consistent with the data-generating mechanism (DGM), we fitted PH regression models for each transition intensity *i.e.* we fitted models of the form: 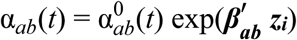, where 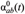 represents the baseline intensity at time *t* when moving from state *a* to state *b*, ***β_ab_*** is the vector of regression parameters, and ***zi*** are the set of (time-fixed) covariates for patient *i*.

We fitted both Cox and Weibull models. We fitted Cox models because they are commonly used in practice. In case of bias due to mis-specification of the baseline intensity function (because it is estimated non-parametrically in the Cox model), we also fitted Weibull PH models (*i.e.* using the same form for the baseline intensity as the DGM).

In our analysis, the estimands of interest were:

i. The vector of transition intensity regression parameters ***βab*** for all possible states *a* and *b*.
ii. The restricted expected length of stay (RELOS) in each state,^4^ restricted to the time period between transplant and two years post-transplant, was used as a summary of the transition probability distributions. RELOS from time 0 to time *t* for state *b* is defined as:

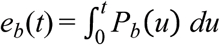 where the state occupation probability, *P_b_*(*t*), denotes the probability of being in state *b* at time *t.*^4^ If all patients are in state 0 initially, *P_b_*(*t*) is equivalent to the transition probability from state 0 to state *b* at time *t, i.e. P_b_*(*t*) *= P*_0*b*_(0*,t*).^20^ We calculated *e_b_*(*t*) using the consistent estimator^4^:

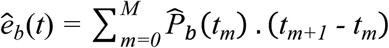 where *P̂*_b_(*t_m_*) is the estimated state occupation probability for state *b* at time *t* and *t_0_< t_1_ < … < t_M_* ≤ *t_M+1_* are the set of ordered times from time 0 up to time *t*, across all transitions. For Cox models, the set of times was the set of all simulated times for the *k^th^* simulation. For Weibull models, the set of times was specified as the set of all values of *t* from time 0 up to time *t*, in increments of 0.1 days.
iii. The final estimand of interest, regression parameter *γ*_12_, was used to test whether our MI approach led to conclusions about the Markov assumption that were consistent with the DGM.^26^ In this test, time from transplant until aGvHD, denoted by *d*, was included as an additional covariate in the model for α_12_ (for the transition from aGvHD to relapse/death), *i.e.* we fitted the model: 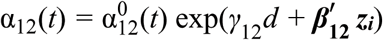. Since our model is Markovian, in truth, *γ*_12_ equals zero.

Performance measures for regression parameters ***β_ab_*** and RELOS were standardised bias (defined as bias/SD of the per-simulation estimates) and average model-based SE. The performance measure of interest for the regression parameter *γ*_12_ was the coverage of the 95% confidence interval *i.e.* the percentage of within-simulation 95% confidence intervals for *γ̂*_12_ that included the true value.

Model-based SE of the regression parameter estimates was calculated using standard methods.^27, 28^ We calculated the model-based SE of RELOS for Cox models using a non-parametric bootstrap estimator,^29^ with 50 bootstraps per simulated dataset, and the model-based SE of RELOS for Weibull models using the delta method. Further details of a separate simulation study to identify the best estimator of model-based SE for Cox and Weibull models are provided in the Supplementary Material (Section S2). The true values of the regression parameters were as per the DGM. The true values of RELOS were calculated using numerical integration.

### 4.3. MI approaches

We considered four MI approaches in this study. Approaches (i) – (iii) did not explicitly account for the ordered nature of the event times. Approaches (ii) - (iv) allowed for different distributions of event times for those who experienced aGvHD compared with those who did not experience aGvHD before relapse/death:

i. “Type 1”^19^ predictive mean matching (PMM), fitting a single imputation model for all patients.
ii. PMM, applying separate imputation models for patients who did and did not experience aGvHD before relapse/death (PMMSUBGP).
iii. MI using draws from a linear imputation model, applying separate imputation models for patients who did and did not experience aGvHD before relapse/death (LINMI). Any negative imputed times were replaced by the value 0.0001 post-imputation.
iv. PMM, compatible with the ordered nature of the event times as specified in the analysis model (PMMCOMP). In this method, PMM was applied using separate imputation models for patients who did and did not experience aGvHD before relapse/death. This method proceeds as follows:

a. Impute the first event time, *i.e.* impute event times to relapse/death for the subgroup of patients who did not experience aGvHD, and event times to aGvHD for the subgroup who did.
b. For the latter group, also impute the (calculated) time from aGvHD to relapse/death, including the time to aGvHD and time from aGvHD to relapse/death in the imputation model, but not the time to relapse/death. Post-imputation, calculate any missing times to relapse/death as the sum of the (observed or imputed) time to aGvHD and the (observed or imputed) time from aGvHD to relapse/death.

Methods (iii) and (iv) were applied in the MCAR scenario. Due to their relatively poor performance, we did not apply these methods in other scenarios. For comparison purposes, we also performed CCA because this method is often used in practice (and is the default method when there are missing values in most statistical software).

Following current guidelines,^30^ each imputation model included all analysis model variables, *i.e.* an indicator of the associated transition (whether times were missing for 0 → 1, 0 → 2, and/or 1 → 2 transitions), both analysis model covariates, indicators of the other events experienced, and associated event times. We used default settings for the number of imputations, iterations, and size of the donor pool for PMM (five in each case) because we wanted to assess the application of MI as routinely used in practice. We concluded that default settings would not unduly influence our results, because we were repeating the analysis for 1000 simulated datasets (which controlled the overall Monte Carlo error even when using a small number of imputations). In addition, in missing data scenarios in which only one variable (the time of aGvHD or relapse/death) was incomplete, no iteration was required. In scenarios in which times to both aGvHD and relapse/death were incomplete, we confirmed that convergence was achieved within five iterations by examining trace plots^31^ for a randomly chosen simulated dataset for each MI method and MDM.

### 4.4. Computer software

Regression parameter estimates and SEs for Cox and Weibull models were calculated using ‘survival’^27^ and ‘flexsurv’^28^ R packages, respectively; state occupation probability estimates were calculated using the ‘mstate’ R package;^32^ MI methods were implemented using the ‘mice’ R package^31^. R code to perform the simulation study is provided in Supplementary Material Section S5.

## 5. Simulation study results

Simulation study results are illustrated in Figures 2 and 3 using “lollipop” plots and all results are included in the Supplementary Material Tables S3a-b. Figures 2 and 3 show the standardised bias of transition intensity regression parameters ***β_ab_*** for each transition, and RELOS within two years, *e_b_*(2), for each state, fitted using a Cox model. Results are illustrated for CCA and the two main MI methods (that is, the methods that were applied in all scenarios): PMM and PMMSUBGP. Bias and model-based SE are not illustrated because these could not be shown on the same scale for all estimands, and because model-based SE was similar for all MI methods and MDMs (and always larger for CCA than for MI methods). Similarly, coverage of the Markov test parameter, *γ_12_*, is not illustrated because with one exception, discussed later, it was similar for all MI methods and MDMs. Figure 2 shows results for scenarios in which MI was expected to work well, that is, when all event times were either MCAR or MAR. Conversely, Figure 3 shows results for scenarios in which MI was not expected to work well, that is, when some or all transition times were MNAR. For comparison, CCA estimates are also illustrated.

**Figure 2.**
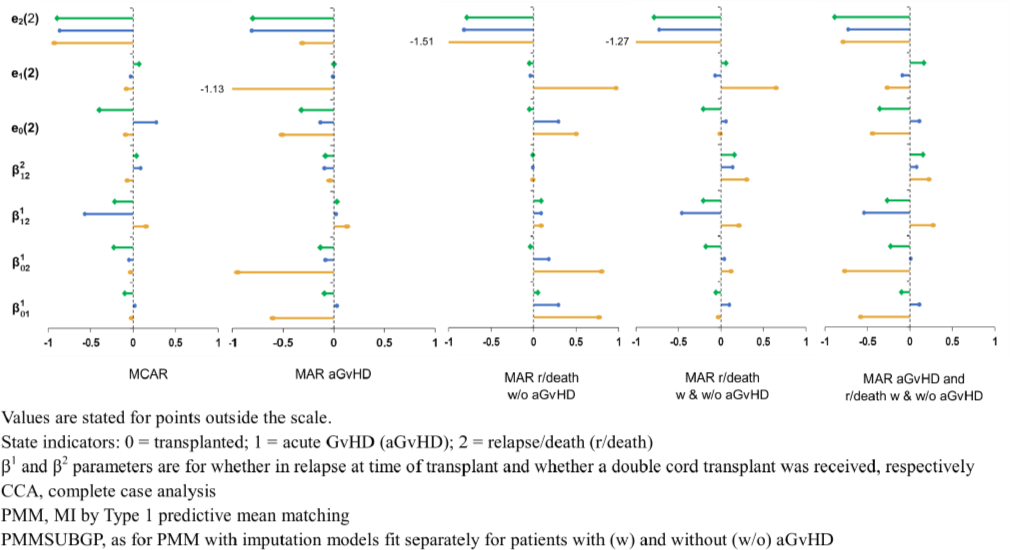
Lollipop plot of standardised bias of transition intensity regression parameters, **βab**, and expected length of stay in each state up to two years post-transplant, e_b_(2), given event times missing completely at random (MCAR) and missing at random (MAR), comparing CCA (yellow oval), PMM (blue circle), and PMMSUBGP (green diamond)

**Figure 3.**
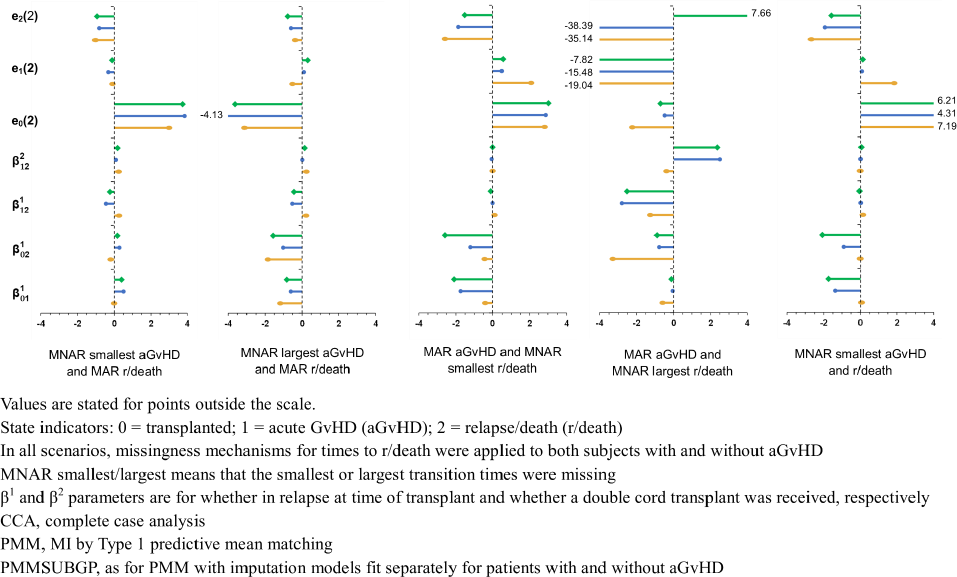
Lollipop plot of standardised bias of transition intensity regression parameters, **β_ab_**, and expected length of stay in each state up to two years post-transplant, e_b_(2), given some event times missing not at random (MNAR), comparing CCA (yellow oval), PMM (blue circle) and PMMSUBGP (green diamond)

As expected, CCA gave unbiased estimates only when event times were MCAR. When event times were either MCAR or MAR (Figure 2), PMM resulted in a small amount of bias for all estimands (that is, the magnitude of the standardised bias was <0.5), except for *e*_2_(2) (RELOS for the relapse/death state) and the regression parameter 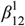 (for the covariate “in relapse or not at time of transplant” in the transition intensity model from aGvHD to relapse/death). The bias in the RELOS estimate, *e*_2_(2), remained for all imputation methods and MDMs when fitting a Cox model, so is not further discussed here. Bias for regression parameter 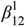 was large in scenarios when event times to relapse/death after aGvHD were missing and small when only event times to aGvHD or relapse/death without aGvHD were missing.

Applying PMM separately for patients who did and did not experience aGvHD before relapse/death (PMMSUBGP) or accounting for the ordered nature of the event times as specified in the analysis model (PMMCOMP) reduced the bias in regression parameter 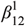. When these methods were used, bias remained small for all other estimands except the RELOS estimate, *e*_2_(2). Results using PMMSUBGP were very similar for both Cox and Weibull models, except that the bias in the RELOS estimate, *e*_2_(2), was greatly reduced when fitting a Weibull model. MI using draws from a linear imputation model (LINMI) resulted in large bias for some estimands, particularly estimates of RELOS (see Supplementary Material for PMMCOMP and LINMI results).

When some or all event times were MNAR (Figure 3), MI using either PMM or PMMSUBGP led to biased estimates. Bias was generally the same or larger than when using CCA. Using MI, bias was larger when the time to the absorbing state (relapse/death) was MNAR than when the time to the intermediate state (aGvHD) was MNAR, and when the largest times were MNAR than when the smallest times were MNAR. Times to relapse/death tended to be longer for patients who experienced aGvHD than for patients who experienced relapse/death without aGvHD (with aGvHD: median 200 days, IQR 386 days; without aGvHD: median 14 days, IQR 22 days). Therefore, MNAR mechanisms where longer times to relapse/death tended to be missing mainly affected patients who experienced aGvHD before relapse/death. Conversely, MNAR mechanisms where shorter times to relapse/death tended to be missing mainly affected patients who experienced relapse/death without aGvHD. This may explain why parameter estimates for the aGvHD to relapse/death transition intensity model, 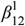 and 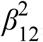, were more biased than parameter estimates for the models of transition from transplant, 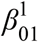 and 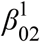, when the largest relapse/death times were MNAR and vice versa when the smallest relapse/death times were MNAR.

As a test of the Markov assumption, the time from transplant until aGvHD was added as a covariate to the transition intensity model from aGvHD to relapse/death. Coverage for the regression parameter for this covariate, *γ*_12_, was in the range 0.92-0.98 in all methods and scenarios, except one. The coverage was 0.66 when applying MI using the PMMSUBGP method and a Weibull analysis model, with aGvHD times MAR and largest relapse/death times MNAR. To allow further exploration of this outlying value for coverage, performance measures for the regression parameter *γ*_12_ are shown in Table 1, for all scenarios in which times to aGvHD were MAR, times to relapse/death times MNAR and the imputation method was PMMSUBGP.

**Table 1.** Performance measures for estimates of the regression parameter γ_12_ in the transition intensity model from aGvHD to relapse/death when some event times are MNAR

| Estimand | | $\gamma_{12}$ | | | |
| --- | --- | --- | --- | --- | --- |
| Missing data mechanism | Multiple imputation method & analysis model | Bias | Mod SE | Std Bias | Cov |
| MAR (aGvHD) & MNAR (smallest times to relapse/death) | PMMSUBGP, Cox | -0.001 | 0.004 | -0.29 | 0.94 |
|  | PMMSUBGP, Weibull | <0.001 | 0.004 | 0.02 | 0.94 |
| MAR (aGvHD) & MNAR (largest times to relapse/death) | PMMSUBGP, Cox | -0.001 | 0.005 | -0.26 | 0.98 |
|  | PMMSUBGP, Weibull | 0.007 | 0.005 | 1.44 | 0.66 |

ModSE, average model-based SE; StdBias, standardised bias; Cov, coverage
Parameter $\gamma_{12}$ is for the time from transplant until aGvHD
PMMSUBGP, MI by Type 1 predictive mean matching with imputation models fit separately for patients with and without aGvHD

As discussed above, MNAR mechanisms in which smallest times to relapse/death times tended to be missing affected mainly patients who experienced relapse/death without aGvHD. Hence, the regression parameter *γ*_12_ is unbiased with coverage close to the nominal value in this scenario. In MNAR mechanisms in which largest times to relapse/death tended to be missing, there is little bias when fitting a Cox model. However, the model-based SE is larger, which may explain the slight over-coverage in this case. Bias is large when fitting a Weibull model, which may explain the high degree of under-coverage in this case.

## 6. Analysis of the Motivating Example

To illustrate our methods, we present an analysis of the NHS CBB dataset. As per the simulation study, our interest was in estimating transition intensity model parameters and RELOS (we estimated RELOS only within one year because event times were sparse beyond this point). Note that our analysis model represents a very simplified version of the events experienced by patients after HSC transplantation. Hence, our results are not intended to be used for clinical insight.

### 6.1. Methods Analysis Model

We fitted the three-state Markov model used in the simulation study, using a PH regression model for each transition intensity (fitting Cox models for all missing data methods, and additionally fitting Weibull models for PMMSUBGP). Transition intensity models included all clinically relevant baseline (at time of transplant) covariates. We tested the analysis model assumptions as follows:

i. The PH assumption was tested for each transition intensity model using the global test (*i.e.* testing for proportional hazards across all covariates in combination) proposed by Grambsch and Therneau.^33^
ii. As a test of the Markov assumption, an additional model was fitted for the transition from aGvHD to relapse/death, including the time from transplant until acute GvHD as well as all covariates.

## Missing Data Methods

In the NHS CBB dataset, both event times and some covariates were partially observed (see Supplementary Material, Section S4). For simplicity, and illustration purposes only, we assumed all data were MAR (see Curnow *et al.*^12^ for discussion of potential missingness mechanisms for this dataset). Therefore, we applied the fully conditional specification (or “chained equations”) MI method^34^ for multiple variables MAR (using the ‘mice’ R package, as before), which involves specifying a series of univariate imputation models, one for each partially observed variable. Covariate data were imputed using standard methods: binary variables using logistic regression, and categorical variables using multinomial regression models. Missing event times were imputed using the main MI methods used in the simulation study (*i.e.* PMM and PMMSUBGP). Here, the sub-groups used in the PMMSUBGP method were:

i. Patients experiencing both acute and chronic GvHD, or chronic GvHD without aGvHD (N=82).
ii. Patients experiencing aGvHD without chronic GvHD (N=173).
iii. Patients experiencing relapse without GvHD, or neither relapse nor GvHD (N=177).

For each MI method, the imputation model for each partially observed variable included all other analysis variables, *i.e.* covariates, an indicator of whether the patient experienced relapse and/or death, and times of aGvHD, relapse, and death. Note that indicators of whether the patient experienced aGvHD or cGvHD were excluded from each imputation model because these had the same value for all patients in each sub-group. Year and country of transplant, and time of cGvHD were also included in each imputation model as auxiliary variables because they were highly predictive of missingness, as well as of the incomplete variables themselves. The time of the composite event (relapse/death) was derived post-imputation.

It is well-established that, to ensure compatibility (or approximate compatibility) with a survival analysis model, both event indicators (binary variables indicating whether each event was experienced) and a representation of the distribution of the associated event times should be included in the imputation model for each partially observed covariate.^35–37^ Since both covariate data and event times were missing in the NHS CBB dataset, the actual event times were included in the imputation models, rather than, for example, the baseline hazard function recommended by White and Royston.^36^ We performed 80 imputations (following the “rule of thumb”^38^ that the number of imputations should at least equal the percentage of incomplete cases - 73% in the NHS CBB dataset). As in the simulation study, we used the default of five iterations per imputation (assessing convergence using trace plots as before) and a donor pool of five donors for each PMM method. We also calculated CCA estimates for comparison purposes.

### 6.2. Results

To illustrate the difference between estimates from CCA and MI methods (PMM and PMMSUBGP, fitting either a Cox or Weibull model for the latter method), Figure 4 shows estimated hazard ratios (HR, conditional on all other covariates) for a double cord transplant (vs. single cord) and whether a patient was in relapse at time of transplant (vs. in remission) for each transition (see Supplementary Material Tables S4a-c for full results). For each HR, 95% confidence intervals (CI) were wider for CCA estimates than for MI estimates. PMMSUBGP estimates were very similar, whether a Cox or Weibull method was fitted. PMM estimates were generally similar to PMMSUBGP estimates. Some CCA point estimates were outside the 95% CI for the equivalent MI estimates (this was the case for double cord transplant, for both the transition from transplant to aGvHD and the transition from aGvHD to relapse/death, and, for some MI estimates, for whether in relapse at time of transplant, for both the transition from transplant to relapse/death and the transition from aGvHD to relapse/death). In these cases, the MI estimates were closer to the null than the CCA estimate.

**Figure 4.**
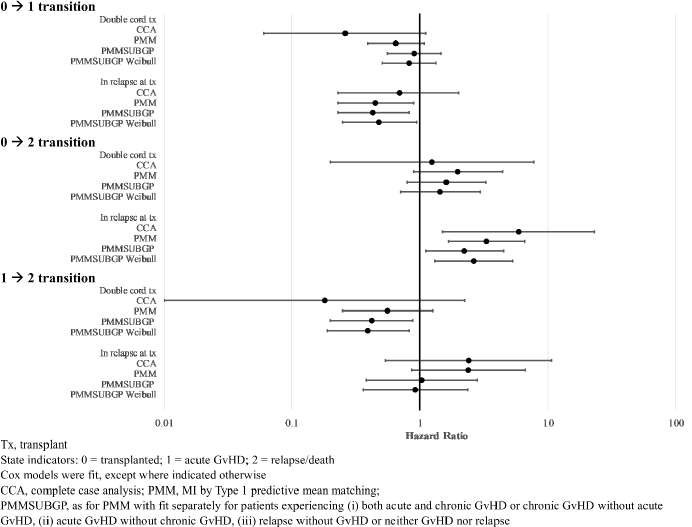
Hazard ratio estimates and 95% confidence intervals for each transition, comparing CCA and MI methods

Table 2 shows CCA and MI estimates of RELOS in the first year post-transplant (illustrated for three different patient types: a patient with reference values of covariates, a low-risk, and a high-risk patient). For each patient type, CCA estimates of the time spent in relapse/death were higher than the MI estimates whilst CCA estimates of time spent in the aGvHD state were lower. For all estimates, CIs were wide and widest for CCA estimates. CIs were also wide for the aGvHD and relapse/death states when fitting a Weibull model (PMMSUBGP Weibull). This may be due to the small number of transitions relative to the range of observed event times *e.g.* only 46 patients with reference values of covariates experienced relapse/death after acute GvHD, and the range of event times was 14 - 1711 days post-transplant. As a consequence, convergence of the Weibull model was not achieved in 29 of the 80 imputed datasets for the transition from aGvHD to relapse/death. There was no apparent association between time of acute GvHD and the hazard of relapse/death after acute GvHD (HR 1.00, 95% CI 0.99-1.01, see Supplementary Material), suggesting there was no violation of the Markov assumption. There was some indication of a violation of the PH assumption, particularly for the model for the transition from transplant to relapse/death (global PH test p-value = 0.14, 0.03, 0.30 for the transitions from transplant to acute GvHD, transplant to relapse/death and acute GvHD to relapse/death, respectively).

**Table 2.** Estimates and 95% confidence intervals (CI) of expected length of stay in each state in the first year post-transplant, comparing CCA and MI methods

| State | Expected length of stay in each state (days) |  |  |  |  |  |  |  |
| --- | --- | --- | --- | --- | --- | --- | --- | --- |
|  | CCA<br>(N=116) |  | PMM<br>(N=432) |  | PMMSUBGP<br>(N=432) |  | PMMSUBGP<br>Weibull (N=432) |  |
|  | Est | 95% CI | Est | 95% CI | Est | 95% CI | Est | 95% CI |
| **Reference patient** |  |  |  |  |  |  |  |  |
| Transplant | 151 | 35-267 | 174 | 111-237 | 158 | 96-220 | 219 | 106-332 |
| AGvHD | 76 | 0-173 | 113 | 56-170 | 132 | 71-193 | 95 | 0-196 |
| Relapse/death | 123 | 0-271 | 70 | 22-118 | 68 | 23-113 | 51 | 0-130 |
| **Low-risk patient** |  |  |  |  |  |  |  |  |
| Transplant | 273 | 0-365 | 229 | 151-307 | 233 | 160-306 | 291 | 212-370 |
| AGvHD | 38 | 0-365 | 95 | 24-166 | 94 | 25-163 | 54 | 0-123 |
| Relapse/death | 39 | 0-365 | 33 | 0-79 | 31 | 0-71 | 20 | 0-61 |
| **High-risk patient** |  |  |  |  |  |  |  |  |
| Transplant | 33 | 0-365 | 72 | 0-167 | 139 | 25-253 | 165 | 0-335 |
| AGvHD | 9 | 0-365 | 22 | 0-54 | 49 | 0-100 | 36 | 0-93 |
| Relapse/death | 309 | 0-365 | 262 | 155-369 | 170 | 45-295 | 163 | 0-360 |

95% CI boundaries outside the range (0,365) were truncated to 0 and 365 for lower and upper bounds respectively.
Unless otherwise stated, Cox transition intensity models were fitted.
CCA, complete case analysis; PMM, MI by Type 1 predictive mean matching;
PMMSUBGP, as for PMM with imputation models fit separately for patients experiencing (i) both acute and chronic GvHD or chronic GvHD without acute GvHD, (ii) acute GvHD without chronic GvHD, (iii) relapse without GvHD or neither GvHD nor relapse.
Reference patient has reference values of all categorical covariates and age 24 years.
Low-risk patient has a non-malignant disorder, receives a high stem cell dose, age 4 years, with reference values of all other covariates.
High-risk patient is in relapse at transplant, receives a double cord transplant, has 2 or more donor-recipient human leucocyte antigen mismatches (matching is important to avoid graft rejection), age 44 years, with reference values of all other covariates.

## 7. Discussion

In this paper, by simulation, we have shown that MSM analysis using Markov models with an MI strategy based on PMM yields estimates with little or negligible bias when event times are MAR. In our setting, in which the probability that event times are missing depends on the event type (a common occurrence in practice because overall survival status is generally completely reported whereas non-fatal events may not be), CCA is not valid because missingness depends on the analysis outcome. Our simulation study shows that even when CCA estimates are unbiased (*e.g.* when times are MCAR), PMM estimates have better precision than CCA estimates. In PMM, missing values are replaced by sampling at random from a donor pool of patients with observed values who are ‘similar’ to the subject with missing data. In the MSM context, this means that the donor pool tends to contain patients who have experienced the same sequence of events as the incomplete case. Therefore, the original sequence of events can be preserved for the incomplete case, without explicitly specifying the order of events in the imputation process. In both our simulation study and real data application, the distribution of event times differed across sub-groups of patients. In the simulation study, applying PMM separately for sub-groups of patients who did and did not experience the intermediate event (PMMSUBGP) tended to reduce bias and model-based SE, particularly for parameters for the transition from the intermediate to absorbing state (aGvHD to relapse/death). PMMSUBGP also improved coverage in a parameter used to test the Markov assumption (by including time from transplant to aGvHD in the model).

An extension of the PMMSUBGP method, which explicitly preserved the ordering of events by including the aGvHD event time and time from aGvHD to relapse/death in the imputation model, but not the relapse/death event time (PMMCOMP), gave results with comparable bias to PMMSUBGP, but larger model-based SE. Due to the loss of information using this method, with no advantage in terms of bias reduction, we would not recommend this approach.

In our study, MI using draws from a linear imputation model (LINMI) led to more bias than PMM when estimating transition intensity model parameters and RELOS. This may be because this approach could result in an imputed relapse/death time that was smaller than the (observed or imputed) aGvHD time. Hence, LINMI was not compatible with the analysis model and estimates were biased as a consequence.

Overall, we recommend using Type 1 PMM to impute missing event times in a MSM analysis using Markov models, first exploring the distribution of event times for each sub-group of patients with a different path through the MSM. Type 1 PMM should be applied separately for each sub-group of patients with a different distribution of event times. In our simulation study, the distributions of simulated times of relapse/death were very different for patients who did and did not experience aGvHD. In analysis of real data, there may be smaller differences between distributions of event times for different sub-groups of patients, and applying PMM by sub-group may make little difference to the results. Therefore, to assess the sensitivity of results to the imputation method, we recommend performing analysis using both a single imputation model and separate models for each sub-group of patients. Note that sub-groups should be of sufficient size to allow for random donor selection in the PMM procedure.

The PMM strategy described here can only be used if some event times are exactly observed, which may not always be the case. For example, after corneal transplantation, hospitals were asked whether any post-transplant surgery had been performed since the previous follow-up report but were not asked for the date of surgery.^39^ In this example, time of surgery would be missing for all patients. Valid use of PMM would require further data collection to obtain exact event times for a representative sample of patients. If this was not possible, a ML approach could be used instead.

Although PMM performed well in our study, there is still scope for improvement, for example, by development of methods that are explicitly compatible with a MSM analysis. This could be achieved, for example, through an extension of the MAR stacked MI approach of Beesley and Taylor^40^ or the SMC-FCS method^41^ to MSM, particularly when these use parametric models. Alternatively, another method proposed by Beesley and Taylor^42^ could be extended, combining an ML approach with full imputation (Beesley and Taylor use “improper” imputation within their EM algorithm).

Generally, MI techniques that assume MAR are not recommended when data are MNAR. In this study, MI resulted in biased estimates when event times were MNAR or a mixture of MAR and MNAR. Our results suggested that bias was greater when the time to the absorbing state (relapse/death) was MNAR than when the time to the intermediate state (aGvHD) was MNAR. This may be due to the constrained nature of the time to an intermediate event (in an illness-death model, this is bounded by 0 and the time of transition to the absorbing state), which may limit the degree of bias even when event times are MNAR. Conversely, the lack of constraint on the maximum time of transition to an absorbing state, and the different pathways through the MSM to that state (each potentially with a different distribution of event times), may increase the degree of bias. Here, we only considered MSMs with the Markov property. Semi-Markov or non-Markov models may result in greater bias when intermediate state event times are MNAR. Therefore, further work is needed to determine if the conclusions of this research still hold for more complex MSMs. A further, useful, extension of this research would be to consider a range of sample sizes, covariate associations, and event rates.

In our simulation study, parametric analysis models generally performed as well as semi-parametric models. Furthermore, parametric models resulted in less biased estimates of the expected length of stay in state (RELOS) when there were sparse event times. However, regression parameter estimates from parametric models were more biased than estimates from semi-parametric models when event times were MNAR. In addition, in practice, parametric models seemed to be more prone to convergence problems than semi-parametric models (and this may have been the case even if all variables were fully observed because event times were sparsely distributed). Further work is required to determine if this is also the case for flexible parametric models.

In the real data analysis, there was some indication that the proportional hazards assumption did not hold, particularly for the model for the transition from transplant to relapse/death. Therefore, the model could be improved by including time-dependent regression parameters, or by using the dynamic landmarking approach.^43^ In addition, clinical inference would be strengthened, and important clinical questions could be answered, if a more detailed event history was modelled for each patient. However, this does rely on the availability of additional post-transplant data, which will almost certainly include some missing data. A more complex analysis model will increase the complexity of any imputation model and the likelihood of imputation model misspecification.

## Data Availability

R code to perform the simulation study is provided in Supplementary Material Section S5. The real data that support the findings of this study are not publicly available due to privacy restrictions.

## Acknowledgements

We are grateful to NHS Blood and Transplant and Eurocord for supplying the UK NHS CBB data.

## Data Availability Statement

The real data that support the findings of this study are not publicly available due to privacy restrictions.

## References

1. Jackson CH. Multi-State Models for Panel Data: The msm Package for R. Journal of Statistical Software. 2011;38(8):1–28.

2. Fiocco M, Putter H, Van Houwelingen HC. Reduced-rank proportional hazards regression and simulation-based prediction for multi-state models. Statist Med. 2008;27:4340–4358.

3. Joly P, Commenges D, Helmer C, Letenneur L. A penalized likelihood approach for an illness–death model with interval-censored data: application to age-specific incidence of dementia. Biostatistics. 2002;3(3):433–443.

4. Grand MK, Putter H. Regression models for expected length of stay. Stat Med. 2016;35:1178–1192.

5. Crowther MJ, Lambert PC. Parametric multistate survival models: Flexible modelling allowing transition-specific distributions with application to estimating clinically useful measures of effect differences. Stat Med. 2017;36:4719–4742.

6. Winton Centre for Risk and Evidence Communication. Communicating the risks & benefits around transplant surgery. 14 September, 2021. https://wintoncentre.maths.cam.ac.uk/projects/communicating-risks-and-benefits-around-transplant-surgery/

7. De Gruttola V, Lagakos SW. Analysis of Doubly-Censored Survival Data, with Application to AIDS. Biometrics. 1989;45:1–11.

8. Lesaffre E, Komárek A. An overview of methods for interval-censored data with an emphasis on applications in dentistry. Stat Methods Med Res. 2005;14:539–552.

9. Machado RJM, Van den Hout A. Flexible multistate models for interval-censored data: Specification, estimation, and an application to ageing research. Stat Med. 2018;37:1636–1649.

10. Yao R, Ananth CV, Park BY, Pereira L, Plante LA. Obesity and the risk of stillbirth: a population-based cohort study. Am J Obstet Gynecol. 2014;210(5):457.e1-457.e9.

11. Carpenter JR, Smuk M. Missing data: A statistical framework for practice. Biom J. 2021;63(5):915–947.

12. Curnow E, Hughes RA, Birnie K, Crowther MJ, May MT, Tilling K. Multiple imputation strategies for a bounded outcome variable in a competing risks analysis. Stat Med. 2021;40:1917–1929.

13. Apperley J, Masszi T. Graft-versus-host disease. In: Apperley J, Carreras E, Gluckman E, Masszi T, eds. Haematopoietic Stem Cell Transplantation: The EBMT Handbook. European School Hematology; 2012.

14. Machado RJM, Hout Avd, Marra G. Penalised maximum likelihood estimation in multi-state models for interval-censored data. Computational Statistics and Data Analysis. 2021;153:107057.

15. Hughes R, Heron J, Sterne J, Tilling K. Accounting for missing data in statistical analyses: multiple imputation is not always the answer. Int J Epidemiol. 2019:1–11.

16. Vansteelandt S, Carpenter JR, Kenward MG. Analysis of Incomplete Data Using Inverse Probability Weighting and Doubly Robust Estimators. Methodology. 2010;6(1):37–48.

17. Little RJA, Zhang N. Subsample ignorable likelihood for regression analysis with missing data. Journal of the Royal Statistical Society: Series C (Applied Statistics). 2011;60(4):591–605.

18. Rubin DB. Multiple imputation for nonresponse in surveys. Wiley; 1987.

19. Morris TP, White IR, Royston P. Tuning multiple imputation by predictive mean matching and local residual draws. BMC Medical Research Methodology 2014;14(75)

20. Andersen PK, Keiding N. Multi-state models for event history analysis. Stat Methods Med Res. 2002;11:91–115.

21. Datta S, Satten GA. Validity of the Aalen-Johansen estimators of stage occupation probabilities and Nelson–Aalen estimators of integrated transition hazards for non-Markov models. Statistics & Probability Letters. 2001;55(4):403–411.

22. Beyersmann J, Latouche Ae, Buchholz A, Schumacher M. Simulating competing risks data in survival analysis. Statist Med. 2009;28:956–971.

23. Van Rood JJ, Stevens CE, Smits J, Carrier C, Carpenter C, Scaradavou A. Reexposure of cord blood to noninherited maternal HLA antigens improves transplant outcome in hematological malignancies. Proc Natl Acad Sci U S A. 2009;106(47):19952–19957.

24. van den Broek BTA, Page K, Paviglianiti A, et al. Early and late outcomes after cord blood transplantation for pediatric patients with inherited leukodystrophies. Blood Advances. 2018;2(1):49–60.

25. Wagner JE, Eapen M, Carter S, et al. One-Unit versus Two-Unit Cord-Blood Transplantation for Hematologic Cancers. N Engl J Med. 2014;371(18):1685–1694.

26. Putter H, Fiocco M, Geskus RB. Tutorial in biostatistics: Competing risks and multi-state models. Stat Med. 2007;26:2389–2430.

27. Therneau T. A Package for Survival Analysis in R. 19 October, 2020. Accessed 19 October, 2020. https://CRAN.R-project.org/package=survival

28. Jackson C. flexsurv: A Platform for Parametric Survival Modeling in R. Journal of Statistical Software. 2016;70(8):1–33.

29. Efron B. Bootstrap methods: another look at the jackknife. The Annals of Statistics. 1979;7(1):1–26.

30. Lee KJ, Tilling K, Cornish RP, et al. Framework for the Treatment And Reporting of Missing data in Observational Studies: The TARMOS framework. J Clin Epidemiol. 2021;134:79–88.

31. Van Buuren S, Groothuis-Oudshoorn K. mice: Multivariate Imputation by Chained Equations in R. Journal of Statistical Software. 2011;45(3):1–67.

32. de Wreede LC, Fiocco M, Putter H. mstate: An R Package for the Analysis of Competing Risks and Multi-State Models. Journal of Statistical Software. 2011;38(7):1–30.

33. Grambsch PM, Therneau TM. Proportional hazards tests and diagnostics based on weighted residuals. Biometrika. 1994;81(3):515–26.

34. Van Buuren S. Multiple imputation of discrete and continuous data by fully conditional specification. Stat Methods Med Res. 2007;16:219–242.

35. Van Buuren S, Boshuizen HC, Knook DL. Multiple imputation of missing blood pressure covariates in survival analysis. Stat Med. 1999;18: 681—694.

36. White IR, Royston P. Imputing missing covariate values for the Cox model. Stat Med. 2009;28:1982–1998.

37. Bartlett JW, Seaman SR, White IR, Carpenter JR. Multiple imputation of covariates by fully conditional specification: Accommodating the substantive model. Stat Methods Med Res. 2015;24(4):462–487.

38. White IR, Royston P, Wood AM. Multiple imputation using chained equations: Issues and guidance for practice. Statistics in Medicine 2011;30:377–399.

39. Steger B, Curnow E, Cheeseman R, et al. Sequential Bilateral Corneal Transplantation and Graft Survival on behalf of the National Health Service Blood and Transplant Ocular Tissue Advisory Group and contributing ophthalmologists Am J Ophthalmol. 2016;170:50–57.

40. Beesley LJ, Taylor JMG. A stacked approach for chained equations multiple imputation incorporating the substantive model. Biometrics. 2020:1–13.

41. Bartlett JW. smcfcs: Multiple Imputation of Covariates by Substantive Model Compatible Fully Conditional Specification. R package version 1.2.1. 19 September, 2016. Accessed September 19, 2016. https://CRAN.R-project.org/package=smcfcs

42. Beesley LJ, Taylor JMG. EM algorithms for fitting multistate cure models. Biostatistics. 2019;20(3):416–432.

43. van Houwelingen HC, Putter H. Dynamic predicting by landmarking as an alternative for multi-state modeling: an application to acute lymphoid leukemia data. Comparative Study Research Support, Non-U.S. Gov’t. Lifetime Data Anal. 14(4):447–63.

