## Supplementary Material for "Multiple imputation strategies for missing event times in a multi-state model analysis"

### Section S1: Simulation study methods

### 1. Application of the method of Beyersmann *et al.* to generate multi-state model data in the simulation study

Multi-state model (MSM) data can be generated by applying the method of Beyersmann *et al.* to a sequence of competing risks experiments. In our case, the sequence of experiments was as follows:

(i) Transitions from transplant ($\alpha_{\text{01}}$ and $\alpha_{\text{02}}$) comprised the first competing risks experiment.
(ii) The transition from acute GvHD to relapse/death ($\alpha_{\text{12}}$) comprised the second experiment. The second experiment was not performed for patients whose first transition was from transplant to relapse/death.

We applied the method of Beyersmann *et al.* under the following assumptions:

- The time of transplant was known, *i.e.* the time origin was observed for all patients.
- Subsequent events could be unobserved, *i.e.* there was right-censoring. The censoring distribution was assumed to be independent of the event time and state occupied.
- For patients who experienced both aGvHD and relapse/death, we assumed that aGvHD always occurred before relapse/death.
- All transitions had the Markov property.
- Each transition intensity model had a proportional hazards (PH) structure. This meant that the transition intensity, $\text{α}_{\text{ab}}$(*t*)*,* at time *t* since transplant, when moving from state *a* to state *b*, was defined for each patient *i* with time-fixed covariates ***z_i_*** as follows:

$\text{α}_{\text{ab}}$(*t*) = $\text{α}_{\text{ab}}^{\text{0}}$(*t*) exp($\text{β}_{\text{ab}}^{\boldsymbol{'}}$ ***z_i_***), where $\text{α}_{\text{ab}}^{\text{0}}$(*t*) represents the baseline intensity at time *t*.

The data generating method proceeds as follows:

**Step 1.** The transition intensity function $\text{α}_{\text{ab}}$(*t*) is defined for each transition as a function of time *t*.

We assumed a Weibull proportional hazards model for each transition intensity function. Our models were informed by our real data, and are defined as follows:

$\text{α}_{\text{01}}$(*t*) = $\left( \frac{\text{1.5}}{\text{36}} \right)\left( \frac{\text{t}}{\text{36}} \right)^{\text{0.5}}$exp{-0.8$\text{z}_{\text{1}}$}

$\text{α}_{\text{02}}$(*t*) = $\left( \frac{\text{0.9}}{\text{120}} \right)\left( \frac{\text{t}}{\text{120}} \right)^{\text{-0.1}}$exp{1.2$\text{z}_{\text{1}}$}

$\text{α}_{\text{12}}$(*t*) = $\left( \frac{\text{0.8}}{\text{160}} \right)\left( \frac{\text{t}}{\text{160}} \right)^{\text{-0.2}}$exp{1.2$\text{z}_{\text{1}}$ – $\text{z}_{\text{2}}$}

where *t* represents the time in days since transplant, $\text{z}_{\text{1}}$ = 1 for a patient in relapse at time of transplant and 0 otherwise, and $\text{z}_{\text{2}}$ = 1 for a double cord transplant and 0 otherwise.

**Step 2.** Event times are generated from the all-cause intensity function (the sum of all transition intensities)$, \sum_{\text{r}} \text{α}_{\text{ab}}\text{(}\text{t}\text{)}$.

$\sum_{\text{r}} \text{α}_{\text{ab}}\text{(}\text{t}\text{)}$ equalled $\text{α}_{\text{01}}$(*t*) + $\text{α}_{\text{02}}$(*t*) for our first competing risks experiment and $\text{α}_{\text{12}}$(*t*) for the second. We used the “simsurv” R package^1^ to generate event times for the first competing risks experiment because in this case the all-cause intensity function could not be inverted using standard analytical methods. For the second experiment, event times were generated from the conditional survival function to ensure that the simulated time to relapse/death, $\text{t}_{\text{12}}$, was greater than the simulated time to acute GvHD, $\text{t}_{\text{01}}$. Times to relapse/death were generated from this function using cumulative hazard inversion, which proceeds as follows:

(a) The conditional survival function is defined as S($\text{t}_{\text{12}}$|$\text{t}_{\text{01}}$) = $\frac{\text{S(}\text{t}_{\text{12}}\text{)}}{\text{S(}\text{t}_{\text{01}}\text{)}}$

where S(*t*) = exp$\left[ \text{-}\left( \frac{\text{t}}{\text{160}} \right)^{\text{0.8}}\text{exp\{1.2}\text{z}_{\text{1}}\text{ }\text{– }\text{z}_{\text{2}}\text{\}} \right]$ with $\text{z}_{\text{1}}$ and $\text{z}_{\text{2}}$ defined as before (based on the standard result for a Weibull distribution^2^).

(b) A value of *u* is drawn from a Uniform(0,1) distribution.
(c) Using the result that the conditional survival function is uniformly distributed across the range (0,1) gives *u* = $\frac{\text{S(}\text{t}_{\text{12}}\text{)}}{\text{S(}\text{t}_{\text{01}}\text{)}}$ ; substituting the expression for S(*t*) and rearranging allows direct calculation of each $\text{t}_{\text{12}}$ as:

$\text{t}_{\text{12}}$ = 160$\left\{ \frac{\text{-}\text{log}\left[ \text{u }\text{exp}\left[ \text{-}\left( \frac{\text{t}_{\text{01}}}{\text{160}} \right)^{\text{0.8}}\text{exp\{}\text{1.2}\text{z}_{\text{1}}\text{ }\text{– }\text{z}_{\text{2}}\text{\}} \right] \right]}{\text{exp\{}\text{1.2}\text{z}_{\text{1}}\text{ }\text{– }\text{z}_{\text{2}}\text{\}}} \right\}^{\text{1/0.8}}$

**Step 3.** A binomial experiment (or multinomial if *r* > 2) is performed to determine the event associated with each event time, with probability $\frac{\text{α}_{\text{ab}}\text{(}\text{t}\text{)}}{\sum_{\text{r}} \text{α}_{\text{ab}}\text{(}\text{t}\text{)}}$ of each of the *r* events occurring.

For our first competing risks experiment, the probability of entering the acute GvHD state (versus relapse/death) from transplant was $\frac{\text{α}_{\text{01}}\text{(}\text{t}_{\text{0*}}\text{)}}{\text{α}_{\text{01}}\text{(}\text{t}_{\text{0*}}\text{) +}\text{ }\text{α}_{\text{02}}\text{(}\text{t}_{\text{0*}}\text{)}}$ , where $\text{t}_{\text{0*}}$ denotes the simulated event time for the first experiment. There was no need to apply this step in the second experiment because the only possible transition from acute GvHD was to relapse/death.

**Step 4.** Censoring times are generated.

In our study, censoring times between one and five years post-transplant were simulated for each patient by taking random draws from a Uniform(365, 1826) distribution.

**2. Missing Data Mechanisms**

Missing data mechanisms used in the simulation study, assuming event times were either missing at random or missing not at random, are summarised in Table S1 overleaf.

*Table S1. Missingness mechanisms used in the simulation study for scenarios 1-11*

| **Scenario** | **Probability of missing event times** | | |
| --- | --- | --- | --- |
|  | **Time to aGvHD** | **Time to relapse/death without aGvHD** | **Time to relapse/death after aGvHD** |
| 1. Times to aGvHD MAR | 0.2 (1 + $\text{z}_{\text{2i}}$) | 0 | 0 |
| 2. Times to aGvHD MNAR (smallest times missing) | 1 if $\text{t}_{\text{i}\text{1}}$ < $\text{t}_{\text{1(30\%)}}$  0 otherwise | 0 | 0 |
| 3. Times to relapse/death MAR (conditional on aGvHD) | 0 | 0.5 (1 – 0.8 $\text{z}_{\text{2i}}$) | 0 |
| 4. Time to relapse/death MAR (not conditional on aGvHD) | 0 | 0.5 (1 – 0.8 $\text{z}_{\text{2i}}$) | 0.5 (1 – 0.8 $\text{z}_{\text{2i}}$) |
| 5. Time to relapse/death MNAR (smallest times missing) | 0 | 1 if $\text{t}_{\text{i}\text{2}}$ < $\text{t}_{\text{RD(30\%)}}$  0 otherwise | 1 if $\text{t}_{\text{i}\text{3}}$ < $\text{t}_{\text{RD(30\%)}}$  0 otherwise |
| 6. Times to aGvHD MAR & times to relapse/death MAR | 0.2 (1 + $\text{z}_{\text{2i}}$) | 0.5 (1 – 0.8 $\text{z}_{\text{2i}}$) | 0.5 (1 – 0.8 $\text{z}_{\text{2i}}$) |
| 7. Times to aGvHD MNAR (smallest times missing) & times to relapse/death MAR | 1 if $\text{t}_{\text{i}\text{1}}$ < $\text{t}_{\text{1(30\%)}}$  0 otherwise | 0.5 (1 – 0.8 $\text{z}_{\text{2i}}$) | 0.5 (1 – 0.8 $\text{z}_{\text{2i}}$) |
| 8. Times to aGvHD MNAR (largest times missing) & times to relapse/death MAR | 1 if $\text{t}_{\text{i}\text{1}}$ > $\text{t}_{\text{1(70\%)}}$  0 otherwise | 0.5 (1 – 0.8 $\text{z}_{\text{2i}}$) | 0.5 (1 – 0.8 $\text{z}_{\text{2i}}$) |
| 9. Times to aGvHD MAR & times to relapse/death MNAR (smallest times missing) | 0.2 (1 + $\text{z}_{\text{2i}}$) | 1 if $\text{t}_{\text{i}\text{2}}$ < $\text{t}_{\text{RD(30\%)}}$  0 otherwise | 1 if $\text{t}_{\text{i}\text{3}}$ < $\text{t}_{\text{RD(30\%)}}$  0 otherwise |
| 10. Times to aGvHD times MAR & times to relapse/death MNAR (largest times missing) | 0.2 (1 + $\text{z}_{\text{2i}}$) | 1 if $\text{t}_{\text{i}\text{2}}$ > $\text{t}_{\text{RD(70\%)}}$  0 otherwise | 1 if $\text{t}_{\text{i}\text{3}}$ > $\text{t}_{\text{RD(70\%)}}$  0 otherwise |
| 11. Times to aGvHD MNAR (smallest times missing) & times to relapse/death MNAR (smallest times missing) | 1 if $\text{t}_{\text{i}\text{1}}$ < $\text{t}_{\text{1(30\%)}}$  0 otherwise | 1 if $\text{t}_{\text{i}\text{2}}$ < $\text{t}_{\text{RD(30\%)}}$  0 otherwise | 1 if $\text{t}_{\text{i}\text{3}}$ < $\text{t}_{\text{RD(30\%)}}$  0 otherwise |

aGvHD, acute graft-versus-host disease; MAR, missing at random; MNAR, missing not at random

In each scenario, for each patient *i*, $\text{z}_{\text{2i}}$ = 1 for a double cord transplant and 0 otherwise;

$\text{t}_{\text{ij}}$ is the event time for patient *i* to the *j^th^* state (*j* = 1: aGvHD, *j* = 2: relapse/death without aGvHD, *j* = 3: relapse/death after aGvHD);

$\text{t}_{\text{j(p\%)}}$ is the *p^th^* percentile of event times to the *j^th^* state, ordered from smallest to largest;

$\text{t}_{\text{RD(}\text{p}\text{\%)}}$ is the *p*^th^ percentile of all times to relapse/death (regardless of whether aGvHD was experienced or not), ordered from smallest to largest.

### Section S2: Derivation of estimators of the standard error of the restricted expected length of stay in state

The three proposed estimators were based on (i) the method of moments, (ii) bootstrapping and (iii) the delta method.

(i) We derived the first estimator, $\hat{\text{SE}}$_1_[${\hat{\text{e}}}_{\text{b}}$(*s*)], by extending Royston and Parmar’s method of moments approach for the restricted mean all-cause survival time.^3^ Following the steps in Royston and Parmar’s derivation, our argument is as follows:

Define the time of transition from state *b*, *T*, with *X* = min*(T*, *s*) representing the transition time restricted to the time period 0 to *s*.

The method of moments estimator is defined as Var (*X*) = E(*X^2^*) – [E(*X*)]^2^

where

E(*X^2^*) = E(*T^2^*|*T* *≤* *s*) P(*T ≤* s*)* + E(*T^2^*|*T* *>* *s*) P(*T > s*)

Here, P(*T > t*) is the state occupation probability $\text{P}_{\text{b}}^{\text{ }}\left( \text{t} \right)$, the probability of being in state *b* at time *t*, and P(*T ≤ t)* = $\text{1 -} \text{P}_{\text{b}}^{\text{ }}\left( \text{t} \right)$, the probability of being in a state other than *b* at time t. Substituting $\text{P}_{\text{b}}^{\text{ }}\left( \text{t} \right)$ and applying integration by parts for the first term in this expression gives:

E(*X^2^*) = *s^2^* [$\text{1 - }\text{P}_{\text{b}}^{\text{ }}\left( \text{s} \right)]$ - $\int_{\text{0}}^{\text{s}} \text{2}\text{t}\text{ [1 - }\text{P}_{\text{b}}^{\text{ }}\left( \text{t} \right)\text{] }\text{dt}$ + *s^2^* $\text{P}_{\text{b}}^{\text{ }}\left( \text{s} \right)$

= *s^2^* - $\int_{\text{0}}^{\text{s}} \text{2}\text{t}\text{ }\text{dt}$ +$\int_{\text{0}}^{\text{s}} \text{2}\text{t}\text{ }\text{P}_{\text{b}}^{\text{ }}\left( \text{t} \right)\text{dt}$

= $\int_{\text{0}}^{\text{s}} \text{2}\text{t}\text{ }\text{P}_{\text{b}}^{\text{ }}\left( \text{t} \right)\text{dt}$

Since E(*X*) = $\int_{\text{0}}^{\text{s}} \text{P}_{\text{b}}^{\text{ }}\left( \text{t} \right)\text{dt}$,

Var (*X*) = $\int_{\text{0}}^{\text{s}} \text{2}\text{t}\text{ }\text{P}_{\text{b}}^{\text{ }}\left( \text{t} \right)\text{dt}$ - $\left[ \int_{\text{0}}^{\text{s}} \text{P}_{\text{b}}^{\text{ }}\left( \text{t} \right)\text{dt} \right]^{\text{2}}$

For the *k^th^* simulation, $\hat{\text{SE}}$_1_[${\hat{\text{e}}}_{\text{b}}^{\text{k}}$(*s*)] = $\sqrt{\frac{\hat{\text{Var}}\text{(}\text{X}\text{)}}{\text{n}}}$, where *n* is the sample size.

Note that Royston and Parmar assume that the underlying transition intensity models are known, but in our study, these must be estimated.

We adapted the expression for the estimator of ${\hat{\text{e}}}_{\text{b}}^{\text{k}}$(*s*), described in Section 3.5 of the paper, to define an estimator of $\hat{\text{Var}}\text{(}\text{X}\text{)}$ for the *k^th^* simulation as follows:

$\hat{\text{Var}}\text{(}\text{X}\text{)}$ = $\sum_{\text{m=0}}^{\text{M}} {\text{2}\text{t}_{\text{m}}^{\text{k}}. \hat{\text{P}}}_{b}^{\text{k}}\left( \text{t}_{\text{m}}^{\text{k}} \right).\left( \text{t}_{\text{m+1}}^{\text{k}}\text{-}\text{t}_{\text{m}}^{\text{k}} \right)$ - $\left[ \sum_{\text{m=0}}^{\text{M}} {\hat{\text{P}}}_{b}^{\text{k}}\left( \text{t}_{\text{m}}^{\text{k}} \right).\left( \text{t}_{\text{m+1}}^{\text{k}}\text{-}\text{t}_{\text{m}}^{\text{k}} \right) \right]^{\text{2}}$

where, for the *k^th^* simulation, ${\hat{\text{P}}}_{\text{b}}^{\text{k}}\left( \text{t} \right)\text{ }$is the estimated probability of being in state *b* at time *t* and $\text{t}_{\text{0}}^{\text{k}}$*<* $\text{t}_{\text{1}}^{\text{k}}$ *< … <* $\text{t}_{\text{M}}^{\text{k}}$ ≤ $\text{t}_{\text{M+1}}^{\text{k}}$ are the set of ordered times up to time *t* post-transplant, across all transitions.

(ii) The second estimator $\hat{\text{SE}}$_2_[${\hat{\text{e}}}_{\text{b}}$(*s*)] was the non-parametric bootstrap estimator,^4^ using 50 bootstraps per simulated dataset. Bartlett and Hughes^5^ note that it is valid to use bootstrapping after MI provided the imputation and analysis models are congenial, but may lead to incorrect coverage if they are not.
(iii) We derived the third estimator, $\hat{\text{SE}}$_3_[${\hat{\text{e}}}_{\text{b}}$(*s*)], using the delta method.

For the Weibull PH model, we used the fact that ${\hat{\text{e}}}_{\text{b}}$(*s*) is a function of the transition intensity model parameter estimates $\hat{\boldsymbol{\theta}}$, with estimated covariance $\hat{\text{Σ}}$, in the application of the delta method. Hence, for the *k^th^* simulation, we defined $\hat{\text{SE}}$_3_[${\hat{\text{e}}}_{\text{b}}^{\text{k}}$((*s*)] as:

$\hat{\text{SE}}$_3_[${\hat{\text{e}}}_{\text{b}}^{\text{k}}$(*s*)] = $\sqrt{\left( \left. \frac{\text{∂}{\hat{\text{e}}}_{\text{b}}^{\text{k}}\text{(s;}\text{θ}\text{)}}{\text{∂}\left( \text{θ} \right)} \right|_{\text{θ}\text{=}\hat{\text{θ}}} \right)^{\text{T}}\hat{\text{Σ}}\left( \left. \frac{\text{∂}{\hat{\text{e}}}_{\text{b}}^{\text{k}}\text{(s;}\text{θ}\text{)}}{\text{∂}\left( \text{θ} \right)} \right|_{\text{θ}\text{=}\hat{\text{θ}}} \right)\text{ }}$

We estimated the vector of partial derivatives $\frac{\text{∂}{\hat{\text{e}}}_{\text{b}}^{\text{k}}\text{(s;}\text{θ}\text{)}}{\text{∂}\left( \text{θ} \right)}$ by calculating the difference in ${\hat{\text{e}}}_{\text{b}}^{\text{k}}$(*s*) from increasing or decreasing each parameter estimate in turn by a small value $\epsilon$, that is:

$\frac{\text{∂}{\hat{\text{e}}}_{\text{b}}^{\text{k}}\text{(s;}\text{θ}\text{)}}{\text{∂}\left( \text{θ} \right)}$ = $\frac{{\hat{\text{e}}}_{\text{b}}^{\text{k}}\text{(}\text{s;}\hat{\text{θ}}\text{+}\text{ϵ}\text{)}\text{ - }{\hat{\text{e}}}_{\text{b}}^{\text{k}}\text{(}\text{s;}\hat{\text{θ}}\text{ - }\text{ϵ}\text{)}\text{ }}{\text{2ϵ }}$

It was not possible to use the same approach for the Cox PH model because the baseline cumulative hazard function in a Cox model is estimated non-parametrically. Instead, we adapted Collett’s definition of SE for the median all-cause survival time,^6^ as follows:

$\hat{\text{SE}}$_3_[${\hat{\text{e}}}_{\text{b}}^{\text{k}}$(*s*)] = $\frac{\text{1}}{{\hat{\text{P'}}}_{b}^{\text{k}}\text{\{}{\hat{\text{e}}}_{\text{b}}^{\text{k}}\text{(}\text{s}\text{)}\text{\}}}\hat{\text{SE}}\text{[}{\hat{\text{P}}}_{\text{b}}^{\text{k}}\text{\{}{\hat{\text{e}}}_{\text{b}}^{\text{k}}\text{(}\text{s}\text{)\}]}$

The SE of the state probability function at ${\hat{\text{e}}}_{\text{b}}^{\text{k}}$(*s*), $\hat{\text{SE}}\text{[}{\hat{\text{P}}}_{b}^{\text{k}}\text{\{}{\hat{\text{e}}}_{\text{b}}^{\text{k}}\text{(}\text{s}\text{)\}]}\text{,}$ was calculated using the Greenwood-style estimator described by de Wreede *et al.*.^7^ We estimated the derivative ${\hat{\text{P'}}}_{\text{b}}^{\text{k}}\text{\{}{\hat{\text{e}}}_{\text{b}}^{\text{k}}\text{(}\text{s}\text{)\}}$ by calculating a local gradient, such that:

${\hat{\text{P'}}}_{\text{b}}^{\text{k}}\text{\{}{\hat{\text{e}}}_{\text{b}}^{\text{k}}\text{(}\text{s}\text{)\}}\text{ =}$ $\frac{{\hat{\text{P}}}_{\text{b}}^{\text{k}}\text{\{}{\hat{\text{u}}}_{\text{b}}^{\text{k}}\text{(}\text{s}\text{)\}}\text{ -}{\hat{\text{P}}}_{\text{b}}^{\text{k}}\text{\{}{\hat{\text{l}}}_{\text{b}}^{\text{k}}\text{(}\text{s}\text{)\}}\text{ }}{{\hat{\text{u}}}_{\text{b}}^{\text{k}}\text{(}\text{s}\text{)}\text{ - }{\hat{\text{l}}}_{\text{b}}^{\text{k}}\text{(}\text{s}\text{)}}$

where ${\hat{\text{u}}}_{b}^{\text{k}}\text{(}\text{s}\text{)}\text{=}\min\left\{ \text{t}_{\text{i}}^{\text{k}} | \text{t}_{\text{i}}^{\text{k}}\text{ ≥ }{\hat{\text{e}}}_{\text{b}}^{\text{k}}\text{(}\text{s}\text{)}\text{ + ϵ} \right\}$ and ${\hat{\text{l}}}_{\text{b}}^{\text{k}}\text{(}\text{s}\text{)}\text{=}\max\left\{ \text{t}_{\text{i}}^{\text{k}} | \text{t}_{\text{i}}^{\text{k}}\text{ }\text{≤}\text{ }{\hat{\text{e}}}_{\text{b}}^{\text{k}}\text{(}\text{s}\text{)}\text{ - ϵ} \right\}$

for all event times *i* for the *b^th^* event type and small $\epsilon$.

In this study, $\epsilon$ was chosen to be 0.00001 for Weibull models and 10 days for Cox models. A large value of $\epsilon$ was needed for the Cox models to accommodate sparse event times; $\epsilon$ needed to be large to ensure that ${\hat{\text{u}}}_{\text{b}}^{\text{k}}\text{(}\text{s}\text{)}$ was different from ${\hat{\text{l}}}_{\text{b}}^{\text{k}}\text{(}\text{s}\text{)}$ for each simulated dataset.

**Methods: study to compare estimators of the standard error of the restricted expected length of stay in state**

The three proposed estimators of the model-based SE of RELOS were compared using a simulation study. Using each of the three proposed estimators, SE of RELOS was estimated for each state *b*, for the time-period between 0 and 2 years post-transplant, using complete data for patients in the 1000 simulated datasets, described previously, with reference values of covariates. Uncensored event times were used so that any differences in SE estimates could not be attributed to increased variation due to censoring. Markov transition intensity models were fitted, assuming, firstly, a Cox PH model and, secondly, a Weibull PH model. Performance measures of interest were the average within-simulation model-based SE and coverage of each estimator of SE. For reference, bias and empirical SE for the estimates of RELOS were also calculated.

The true values of *e_b_*(2) for each state *j* were calculated as $\int_{\text{0}}^{\text{2}} \text{P}_{\text{b}}\left( \text{t} \right)\text{ dt}$, using numerical integration. The transition intensity models specified in the DGM were substituted into standard expressions for $\text{P}_{\text{0}}\left( \text{t} \right)$, $\text{P}_{\text{1}}\left( \text{t} \right)\text{ }$and $\text{P}_{\text{2}}\left( \text{t} \right)\text{ }$for a Markov illness-death model,^8^ namely:

$\text{P}_{\text{0}}\left( \text{t} \right)\text{ }$*=* exp $\left\{ \text{-}\int_{\text{0}}^{\text{t}} \text{α}_{\text{01}}\text{(}\text{s}\text{)}\text{+ }\text{α}_{\text{02}}\text{(}\text{s}\text{)}\text{ ds} \right\}$

$\text{P}_{\text{1}}\left( \text{t} \right)$*=* $\int_{\text{0}}^{\text{t}} \text{α}_{\text{01}}\text{(}\text{s}\text{)}\text{ }\text{exp}\left\{ \text{-}\int_{\text{s}}^{\text{t}} \text{α}_{\text{12}}\text{(}\text{u}\text{)}\text{du} \right\}\text{P}_{\text{0}}\text{(}\text{s}\text{)}\text{ ds}$

$\text{P}_{\text{2}}\left( \text{t} \right)$ *=* $\int_{\text{0}}^{\text{t}} \left( \text{α}_{\text{01}}\text{(}\text{s}\text{)}\left( \text{1}\text{ – }\text{exp}\left\{ \text{-}\int_{\text{s}}^{\text{t}} \text{α}_{\text{12}}\text{(}\text{u}\text{)}\text{du} \right\} \right)\text{ + }\text{α}_{\text{02}}\text{(}\text{s}\text{)} \right).\text{P}_{\text{0}}\text{(}\text{s}\text{)}\text{ ds}$

In addition, the theoretical values of the extended Royston and Parmar estimator, $\text{SE}_{\text{1}}\text{[}{\hat{\text{e}}}_{\text{b}}\text{(}\text{s}\text{)]}$, were calculated using the same method.

### Results: study to compare estimators of the standard error of the restricted expected length of stay in state

The average within-simulation estimated model-based SE and coverage of the three estimators of the SE of ${\hat{\text{e}}}_{\text{b}}\text{(2)}$ for each state *b*, fitting either a Cox or Weibull model, are shown in Table S2. For reference, bias and empirical SE of the estimates of ${\hat{\text{e}}}_{\text{b}}\text{(2)}$, and theoretical (analytical) values of the extended Royston and Parmar estimator, $\text{SE}_{\text{1}}\text{[}{\hat{\text{e}}}_{\text{b}}\text{(2)]}$, are also shown.

*Table S2. Average model-based standard error (ModSE) and coverage (Cov) of each estimator of the SE of RELOS between 0 and 2 years post-transplant*

| **Model** | **State** | **Bias of**  ${\hat{\text{e}}}_{\text{b}}\text{(2)}$ | **EmpSE of**  ${\hat{\text{e}}}_{\text{b}}\text{(2}$**)** | **Estimator of SE of** ${\hat{\text{e}}}_{\text{b}}\text{(2)}$ | | | | | | |
| --- | --- | --- | --- | --- | --- | --- | --- | --- | --- | --- |
|  |  |  |  | **Royston & Parmar** | | | **Bootstrap** | | **Delta** | |
|  |  |  |  | **TheorSE** | **ModSE** | **Cov** | **ModSE** | **Cov** | **ModSE** | **Cov** |
| Cox | Tx | -0.05 | 1.12 | 0.89 | 0.94 | 0.90 | 1.42 | 0.96 | 1.32 | 0.97 |
|  | AgvHD | -0.38 | 11.12 | 9.13 | 9.00 | 0.90 | 11.35 | 0.95 | 17.10 | 0.98 |
|  | R/death | -10.00 | 15.17 | 17.51 | 16.98 | 0.93 | 15.90 | 0.87 | 111.60 | >0.99 |
| Weibull | Tx | 0.02 | 1.04 | 0.89 | 0.88 | 0.91 | 0.99 | 0.92 | 0.99 | 0.94 |
|  | AgvHD | -0.18 | 11.04 | 9.13 | 9.10 | 0.90 | 11.24 | 0.94 | 11.26 | 0.95 |
|  | R/death | 0.16 | 11.18 | 17.51 | 17.50 | >0.99 | 11.37 | 0.95 | 11.40 | 0.95 |

${\hat{\text{e}}}_{\text{b}}\text{(2)}$, RELOS between 0 and 2 years post-transplant for state *b*; EmpSE, empirical SE; TheorSE, theoretical SE for Royston & Parmar estimator; Tx, transplant; AgvHD, acute GvHD; R/death, relapse/death.

Estimates use uncensored event times for patients with reference values of the covariates, based on 1000 simulated datasets.

Monte Carlo SE for bias was 0.04, 0.03 for ${\hat{\text{e}}}_{\text{0}}\text{(2)}$**,** 0.35, 0.35 for ${\hat{\text{e}}}_{\text{1}}\text{(2)}$**,** and 0.48, 0.35 for ${\hat{\text{e}}}_{\text{2}}\text{(2)}$ for Cox and Weibull models, respectively.

Monte Carlo SE for coverage was ≤ 0.01 in all cases.

The theoretical (analytical) values of RELOS between 0 and 2 years post-transplant were$\text{ }\text{e}_{\text{0}}\text{(2)}$ **=** 26.3 days, $\text{e}_{\text{1}}\text{(2)}$ **=** 140.6 days, and $\text{e}_{\text{2}}\text{(2)}$ **=** 563.1 days. Values of RELOS estimated from the simulated data were unbiased for both Cox and Weibull models, except for the relapse/death state when fitting a Cox model (given Monte Carlo 95% CI for bias of (-0.13, 0.03) for the transplant state, (-1.07, 0.31) for the acute GvHD state, and (-10.94, -9.06) for the relapse/death state for Cox models; and (-0.04, 0.08) for the transplant state, (-0.87, 0.51) for the acute GvHD state, and (-0.53, 0.85) for the relapse/death state for Weibull models). The bias in this case can be explained by the large time intervals between individual simulated relapse/death event times.

Estimates of SE using the extended Royston and Parmar (R&P) method, $\hat{\text{SE}}$_1_[${\hat{\text{e}}}_{\text{j}}$(*s*)], were very similar to the theoretical calculated values for this estimator, for both Cox and Weibull models. However, the R&P estimated SEs differed from the empirical SEs, as follows. For the transplant and acute GvHD states, R&P estimated SEs were less than empirical SE and, for the relapse/death state, R&P estimated SEs were greater than empirical SE. The difference between the R&P estimated SEs and empirical SE here may be because the Royston and Parmar method treats model parameters as fixed. Hence, the covariance associated with model parameter estimation (which may have a positive or negative effect on SE) is not accounted for. The under-estimation of the SE could explain the under-coverage for transplant and acute GvHD states for both Cox and Weibull models (since the point estimates are unbiased).^9^ Similarly, the over-estimation of the SE could explain the over-coverage for the relapse/death state for the Weibull model. The bias in the estimate of RELOS for the relapse/death state for the Cox model could explain the under-coverage in this case.^9^

Estimates of SE using the bootstrap method, $\hat{\text{SE}}$_2_[${\hat{\text{e}}}_{\text{j}}$(*s*)], were very similar to the empirical SE for all three states, for both Cox and Weibull models. However, there was under-coverage for the relapse/death state from the Cox model, and for the transplant state from the Weibull model. As before, the bias in the estimate of RELOS for the relapse/death state from the Cox model explains the greater degree of under-coverage in this case. For the Weibull model, the bootstrap estimated SE is slightly lower than the empirical SE for the transplant state, which explains the under-coverage in this case.

Estimates of SE using the delta method, $\hat{\text{SE}}$_3_[${\hat{\text{e}}}_{\text{j}}$(*s*)], were also very similar to the empirical SE for all three states from Weibull models, with appropriate coverage. However, for the Cox models, performance of this estimator was poor, with over-coverage for all states. For the transplant state, the delta estimated SE was similar to the empirical SE. However, for acute GvHD and relapse/death states, delta estimated SEs were larger than the empirical SEs; for the relapse/death state, the SE estimate was nearly 10 times larger. For the acute GvHD state, this can be explained by the implicit assumption of this method that all patients will be in each state at some time *t* > 0. This is true for the initial and absorbing states, but not necessarily for the intermediate state (some patients will relapse/die without experiencing acute GvHD). For the relapse/death state, this can be explained by the large time intervals between individual simulated relapse/death event times; Royston and Parmar^3^ comment that the delta method does not work well with sparse event times. The results for the bootstrap and delta method (as applied to the Weibull model) suggest that, in contrast to the R&P method, both these methods appropriately incorporate the covariance associated with model parameter estimation. The best-performing estimators of the SE of RELOS were used in the main simulation study. The bootstrap estimator was used when fitting Cox models and the delta estimator was used when fitting Weibull models.

**References for Sections S1 and S2**

1.	Brilleman SL. simsurv: Simulate Survival Data. 10 February, 2020. Accessed 10 February, 2020. <https://CRAN.R-project.org/package=simsurv>

2.	Bender R, Augustin T, Blettner M. Generating survival times to simulate Cox proportional hazards models. *Stat Med*. 2005;24:1713-1723.

3.	Royston P, Parmar MK. Restricted mean survival time: an alternative to the hazard ratio for the design and analysis of randomized trials with a time-to-event outcome. *BMCMedical Research Methodology*. 2013;13(152):1-15.

4.	Efron B. Bootstrap methods: another look at the jackknife. *The Annals of Statistics*. 1979;7(1):1-26.

5.	Bartlett JW, Hughes RA. Bootstrap inference for multiple imputation under uncongeniality and misspecification. *Stat Methods Med Res*. 2020;29(12):3533-3546.

6.	Collett D. *Modelling Survival Data in Medical Research* 3rd ed. Chapman and Hall /CRC; 2015.

7.	Wreede LCd, Fiocco M, Putter H. The mstate package for estimation and prediction in non- and semi-parametric multi-state and competing risks models. *Computer Methods and Programs in Biomedicine* 2010;99:261–274.

8.	Putter H, Fiocco M, Geskus RB. Tutorial in biostatistics: Competing risks and multi-state models. *Stat Med*. 2007;26:2389–2430.

9.	Morris TP, White IR, Crowther MJ. Using simulation studies to evaluate statistical methods. *Stat Med*. 2019;38:2074-2102.

**Section S3: Simulation Study Results**

Table S3a. Simulation study results. Bias, average model-based SE (ModSE), empirical SE (EmpSE) and standardised bias (StdBias) of regression parameters **β**_ab_ and coverage (Cov) for regression parameter γ_12_ for each transition intensity model, given various missing data mechanisms and analysis approaches.

| **Estimand (true result)** | | $\text{β}_{\text{01}}^{\text{1}}$ **(-0.8)** | | | | $\text{β}_{\text{02}}^{\text{1}}$ **(1.2)** | | | | $\text{β}_{\text{12}}^{\text{1}}$ **(1.2)** | | | | $\text{β}_{\text{12}}^{\text{2}}$ **(-1.0)** | | | | $\text{γ}_{\text{12}}$**(0)** |
| --- | --- | --- | --- | --- | --- | --- | --- | --- | --- | --- | --- | --- | --- | --- | --- | --- | --- | --- |
| **Missing data mechanism** | **Analysis approach** | **Bias** | **Mod**  **SE** | **Emp**  **SE** | **Std**  **Bias** | **Bias** | **Mod**  **SE** | **Emp**  **SE** | **Std**  **Bias** | **Bias** | **Mod**  **SE** | **Emp**  **SE** | **Std**  **Bias** | **Bias** | **Mod**  **SE** | **Emp**  **SE** | **Std**  **Bias** | **Cov** |
| **Complete data (Cox)** | | 0.00 | 0.20 | 0.20 | 0.02 | 0.00 | 0.16 | 0.16 | -0.01 | 0.02 | 0.21 | 0.21 | 0.09 | 0.00 | 0.13 | 0.13 | -0.01 | 0.94 |
| **Complete data (Weibull)** | | 0.00 | 0.20 | 0.20 | 0.01 | 0.00 | 0.16 | 0.16 | 0.02 | 0.02 | 0.21 | 0.21 | 0.11 | -0.01 | 0.13 | 0.13 | -0.04 | 0.95 |
| **MCAR** | CCA | -0.01 | 0.29 | 0.30 | -0.02 | -0.01 | 0.23 | 0.23 | -0.03 | 0.05 | 0.31 | 0.30 | 0.15 | -0.01 | 0.18 | 0.19 | -0.07 | 0.95 |
|  | PMM | 0.00 | 0.25 | 0.21 | 0.02 | -0.01 | 0.19 | 0.16 | -0.05 | -0.13 | 0.31 | 0.23 | -0.57 | 0.01 | 0.16 | 0.15 | 0.09 | 0.95 |
|  | PMMSUBGP | -0.02 | 0.24 | 0.23 | -0.10 | -0.04 | 0.18 | 0.17 | -0.23 | -0.06 | 0.29 | 0.26 | -0.22 | 0.01 | 0.16 | 0.15 | 0.04 | 0.94 |
|  | PMMSUBGP* | -0.03 | 0.24 | 0.23 | -0.12 | -0.03 | 0.18 | 0.16 | -0.17 | -0.05 | 0.29 | 0.25 | -0.18 | -0.02 | 0.16 | 0.15 | -0.11 | 0.95 |
|  | PMMCOMP | 0.02 | 0.23 | 0.22 | 0.09 | -0.01 | 0.17 | 0.17 | -0.03 | -0.07 | 0.37 | 0.32 | -0.22 | -0.01 | 0.20 | 0.20 | -0.06 | 0.96 |
|  | LINMI | 0.00 | 0.26 | 0.21 | 0.02 | -0.11 | 0.20 | 0.15 | -0.68 | -0.15 | 0.32 | 0.24 | -0.64 | 0.05 | 0.17 | 0.15 | 0.31 | 0.95 |
| **MAR (acute GvHD only)** | CCA | -0.15 | 0.24 | 0.25 | -0.60 | -0.15 | 0.16 | 0.16 | -0.95 | 0.03 | 0.25 | 0.25 | 0.13 | -0.01 | 0.16 | 0.16 | -0.04 | 0.93 |
|  | PMM | 0.01 | 0.22 | 0.20 | 0.03 | -0.01 | 0.16 | 0.16 | -0.08 | 0.01 | 0.21 | 0.21 | 0.02 | -0.01 | 0.13 | 0.13 | -0.09 | 0.97 |
|  | PMMSUBGP | -0.02 | 0.22 | 0.22 | -0.09 | -0.02 | 0.16 | 0.16 | -0.13 | 0.01 | 0.21 | 0.21 | 0.03 | -0.01 | 0.13 | 0.13 | -0.08 | 0.94 |
|  | PMMSUBGP* | -0.02 | 0.22 | 0.22 | -0.08 | -0.01 | 0.16 | 0.16 | -0.08 | 0.01 | 0.21 | 0.21 | 0.05 | -0.02 | 0.13 | 0.13 | -0.16 | 0.95 |
| **MNAR (smallest acute GvHD only)** | CCA | 0.00 | 0.24 | 0.24 | 0.01 | -0.06 | 0.16 | 0.16 | -0.39 | 0.03 | 0.25 | 0.25 | 0.14 | 0.00 | 0.15 | 0.16 | -0.01 | 0.94 |
|  | PMM | 0.11 | 0.23 | 0.22 | 0.51 | 0.04 | 0.16 | 0.15 | 0.26 | 0.02 | 0.22 | 0.22 | 0.09 | 0.00 | 0.13 | 0.13 | 0.00 | 0.96 |
|  | PMMSUBGP | 0.09 | 0.23 | 0.23 | 0.38 | 0.04 | 0.15 | 0.15 | 0.23 | 0.02 | 0.22 | 0.22 | 0.10 | 0.00 | 0.13 | 0.13 | 0.00 | 0.96 |
|  | PMMSUBGP* | 0.03 | 0.23 | 0.26 | 0.11 | 0.04 | 0.15 | 0.15 | 0.27 | 0.02 | 0.22 | 0.22 | 0.11 | 0.00 | 0.13 | 0.13 | -0.02 | 0.95 |
| **MAR (relapse/ death only, 0→2 transition)** | CCA | 0.17 | 0.20 | 0.20 | 0.77 | 0.16 | 0.19 | 0.20 | 0.80 | 0.02 | 0.21 | 0.21 | 0.09 | 0.00 | 0.13 | 0.13 | -0.01 | 0.94 |
|  | PMM | 0.06 | 0.25 | 0.21 | 0.29 | 0.03 | 0.20 | 0.17 | 0.18 | 0.02 | 0.21 | 0.21 | 0.09 | 0.00 | 0.13 | 0.13 | -0.01 | 0.94 |
|  | PMMSUBGP | 0.01 | 0.21 | 0.21 | 0.05 | -0.01 | 0.17 | 0.16 | -0.04 | 0.02 | 0.21 | 0.21 | 0.09 | 0.00 | 0.13 | 0.13 | -0.01 | 0.94 |
|  | PMMSUBGP* | 0.01 | 0.21 | 0.22 | 0.05 | 0.00 | 0.16 | 0.17 | 0.03 | 0.02 | 0.21 | 0.21 | 0.11 | -0.01 | 0.13 | 0.13 | -0.04 | 0.95 |
| **MAR (relapse/ death only, both transitions)** | CCA | -0.01 | 0.24 | 0.25 | -0.03 | 0.02 | 0.19 | 0.19 | 0.12 | 0.06 | 0.26 | 0.27 | 0.21 | 0.05 | 0.15 | 0.16 | 0.30 | 0.94 |
|  | PMM | 0.02 | 0.26 | 0.22 | 0.10 | 0.01 | 0.22 | 0.17 | 0.04 | -0.12 | 0.34 | 0.26 | -0.46 | 0.02 | 0.17 | 0.16 | 0.14 | 0.94 |
|  | PMMSUBGP | -0.01 | 0.22 | 0.22 | -0.06 | -0.03 | 0.18 | 0.17 | -0.18 | -0.06 | 0.31 | 0.28 | -0.21 | 0.03 | 0.16 | 0.16 | 0.16 | 0.94 |
|  | PMMSUBGP* | -0.02 | 0.22 | 0.21 | -0.10 | -0.02 | 0.18 | 0.16 | -0.13 | -0.05 | 0.30 | 0.28 | -0.18 | 0.00 | 0.16 | 0.16 | -0.03 | 0.94 |
| **MNAR (smallest relapse/death only, both transitions)** | CCA | 0.05 | 0.21 | 0.22 | 0.23 | 0.00 | 0.32 | 0.33 | -0.01 | 0.02 | 0.22 | 0.22 | 0.09 | 0.00 | 0.13 | 0.13 | -0.01 | 0.94 |
|  | PMM | -0.37 | 0.26 | 0.22 | -1.69 | -0.30 | 0.44 | 0.25 | -1.21 | 0.01 | 0.23 | 0.22 | 0.07 | 0.00 | 0.13 | 0.13 | -0.02 | 0.95 |
|  | PMMSUBGP | -0.45 | 0.22 | 0.22 | -2.07 | -0.58 | 0.23 | 0.22 | -2.61 | -0.01 | 0.23 | 0.22 | -0.03 | 0.00 | 0.13 | 0.13 | 0.03 | 0.94 |
|  | PMMSUBGP* | -0.45 | 0.22 | 0.22 | -2.10 | -0.57 | 0.25 | 0.29 | -1.93 | 0.01 | 0.22 | 0.22 | 0.05 | -0.04 | 0.13 | 0.13 | -0.27 | 0.92 |
| **MAR (acute GvHD & relapse/death)** | CCA | -0.18 | 0.30 | 0.32 | -0.58 | -0.15 | 0.19 | 0.19 | -0.77 | 0.09 | 0.32 | 0.32 | 0.27 | 0.04 | 0.19 | 0.19 | 0.22 | 0.94 |
|  | PMM | 0.02 | 0.28 | 0.23 | 0.11 | 0.00 | 0.22 | 0.18 | 0.01 | -0.14 | 0.35 | 0.26 | -0.54 | 0.01 | 0.16 | 0.15 | 0.08 | 0.92 |
|  | PMMSUBGP | -0.02 | 0.24 | 0.23 | -0.10 | -0.04 | 0.18 | 0.16 | -0.23 | -0.08 | 0.32 | 0.27 | -0.27 | 0.02 | 0.17 | 0.15 | 0.15 | 0.93 |
|  | PMMSUBGP* | -0.03 | 0.24 | 0.23 | -0.13 | -0.03 | 0.18 | 0.16 | -0.19 | -0.06 | 0.31 | 0.27 | -0.22 | -0.02 | 0.17 | 0.16 | -0.14 | 0.95 |
| **MNAR (smallest acute GvHD) & MAR (relapse/death)** | CCA | 0.00 | 0.29 | 0.29 | -0.01 | -0.04 | 0.19 | 0.20 | -0.20 | 0.08 | 0.31 | 0.30 | 0.26 | 0.05 | 0.18 | 0.19 | 0.24 | 0.94 |
|  | PMM | 0.12 | 0.30 | 0.24 | 0.51 | 0.05 | 0.22 | 0.17 | 0.28 | -0.12 | 0.36 | 0.27 | -0.46 | 0.01 | 0.17 | 0.16 | 0.09 | 0.94 |
|  | PMMSUBGP | 0.10 | 0.26 | 0.24 | 0.40 | 0.03 | 0.17 | 0.16 | 0.17 | -0.07 | 0.33 | 0.29 | -0.24 | 0.03 | 0.17 | 0.15 | 0.18 | 0.96 |
|  | PMMSUBGP* | 0.04 | 0.26 | 0.28 | 0.15 | 0.04 | 0.17 | 0.16 | 0.23 | -0.05 | 0.33 | 0.27 | -0.19 | 0.00 | 0.17 | 0.16 | -0.03 | 0.95 |
| **MNAR (largest acute GvHD) & MAR (relapse/ death)** | CCA | -0.38 | 0.30 | 0.33 | -1.17 | -0.40 | 0.20 | 0.22 | -1.85 | 0.07 | 0.32 | 0.33 | 0.22 | 0.04 | 0.18 | 0.19 | 0.24 | 0.94 |
|  | PMM | -0.15 | 0.28 | 0.25 | -0.59 | -0.21 | 0.27 | 0.21 | -1.01 | -0.14 | 0.35 | 0.27 | -0.51 | 0.00 | 0.17 | 0.15 | 0.01 | 0.97 |
|  | PMMSUBGP | -0.19 | 0.22 | 0.23 | -0.81 | -0.31 | 0.19 | 0.20 | -1.56 | -0.11 | 0.31 | 0.27 | -0.42 | 0.02 | 0.17 | 0.15 | 0.14 | 0.95 |
|  | PMMSUBGP* | -0.31 | 0.23 | 0.26 | -1.20 | -0.21 | 0.18 | 0.17 | -1.24 | -0.11 | 0.30 | 0.26 | -0.42 | -0.03 | 0.17 | 0.16 | -0.16 | 0.95 |
| **MAR (acute GvHD) & MNAR (smallest relapse/death)** | CCA | -0.11 | 0.25 | 0.27 | -0.40 | -0.14 | 0.32 | 0.33 | -0.43 | 0.03 | 0.26 | 0.27 | 0.11 | 0.00 | 0.16 | 0.17 | 0.00 | 0.94 |
|  | PMM | -0.38 | 0.26 | 0.22 | -1.74 | -0.31 | 0.43 | 0.26 | -1.21 | 0.00 | 0.23 | 0.22 | 0.00 | -0.01 | 0.13 | 0.13 | -0.05 | 0.94 |
|  | PMMSUBGP | -0.46 | 0.23 | 0.22 | -2.10 | 0.61 | 0.24 | 0.24 | -2.57 | -0.02 | 0.23 | 0.22 | -0.10 | 0.00 | 0.13 | 0.13 | 0.00 | 0.94 |
|  | PMMSUBGP* | -0.47 | 0.23 | 0.22 | -2.13 | -0.59 | 0.27 | 0.31 | -1.90 | 0.00 | 0.23 | 0.22 | 0.00 | -0.04 | 0.13 | 0.14 | -0.31 | 0.94 |
| **MAR (acute GvHD) & MNAR (largest relapse/death)** | CCA | -0.16 | 0.27 | 0.27 | -0.60 | -0.52 | 0.16 | 0.16 | -3.30 | -0.37 | 0.28 | 0.30 | -1.25 | -0.09 | 0.23 | 0.24 | -0.39 | 0.95 |
|  | PMM | -0.01 | 0.23 | 0.21 | -0.05 | -0.13 | 0.18 | 0.16 | -0.79 | -0.57 | 0.25 | 0.20 | -2.80 | 0.32 | 0.17 | 0.13 | 2.50 | 0.98 |
|  | PMMSUBGP | -0.03 | 0.23 | 0.22 | -0.12 | -0.15 | 0.18 | 0.16 | -0.90 | -0.54 | 0.25 | 0.22 | -2.52 | 0.31 | 0.18 | 0.13 | 2.36 | 0.98 |
|  | PMMSUBGP* | -0.03 | 0.23 | 0.22 | -0.12 | -0.14 | 0.18 | 0.16 | -0.87 | -0.48 | 0.23 | 0.18 | -2.73 | 0.08 | 0.16 | 0.15 | 0.53 | 0.66 |
| **MNAR (smallest acute GvHD & smallest relapse/death)** | CCA | 0.01 | 0.24 | 0.24 | 0.06 | 0.00 | 0.32 | 0.33 | -0.01 | 0.03 | 0.25 | 0.26 | 0.13 | 0.00 | 0.15 | 0.16 | -0.01 | 0.94 |
|  | PMM | -0.31 | 0.28 | 0.23 | -1.37 | -0.22 | 0.39 | 0.24 | -0.90 | 0.00 | 0.24 | 0.22 | 0.02 | 0.00 | 0.13 | 0.13 | 0.01 | 0.94 |
|  | PMMSUBGP | -0.41 | 0.24 | 0.23 | -1.74 | -0.46 | 0.23 | 0.22 | -2.08 | -0.01 | 0.24 | 0.22 | -0.05 | 0.01 | 0.13 | 0.13 | 0.05 | 0.95 |
|  | PMMSUBGP* | -0.44 | 0.26 | 0.26 | -1.67 | -0.45 | 0.25 | 0.29 | -1.54 | 0.00 | 0.23 | 0.22 | -0.01 | -0.01 | 0.13 | 0.13 | -0.06 | 0.95 |

State indicators: 0 = transplanted; 1 = acute GvHD; 2 = relapse/death.

*Cox models were fit, except for methods indicated with a *, for which Weibull models were fit.

Parameters $\beta_{lm}^{1}$, $\beta_{lm}^{2}$, *γ_12_* are for whether in relapse at time of transplant, double cord transplanted and time from transplant until acute GvHD, respectively.

Monte Carlo SE for bias/coverage ranges from 0.004 to 0.015 for all estimands.

CCA, complete case analysis; PMM, MI by Type 1 predictive mean matching; PMMSUBGP, as for PMM with imputation models fit separately for patients with and without acute GvHD; PMMCOMP, as for PMM, imputing time to acute GvHD and time from acute GvHD to relapse/death*,* with post-imputation calculation of time from transplant to relapse/death; LINMI, MI using draws from a linear imputation model, with imputation models fit separately for patients with and without acute GvHD.

Table S3b. Simulation study results. Bias, average model-based SE (ModSE), empirical SE (EmpSE) and standardised bias (StdBias) of RELOS between 0 and 2 years, e_b_(2), for each transition intensity model given various missing data mechanisms and analysis approaches.

| **Estimand**  **(true result)** | | $\text{e}_{\text{0}}\text{(2)}$  **(26.3)** | | | | $\text{e}_{\text{1}}\text{(2)}$  **(140.6)** | | | | $\text{e}_{\text{2}}\text{(2)}$  **(563.1)** | | | |
| --- | --- | --- | --- | --- | --- | --- | --- | --- | --- | --- | --- | --- | --- |
| **Missing data mechanism** | **Analysis approach** | **Bias** | **Mod**  **SE** | **Emp**  **SE** | **Std**  **Bias** | **Bias** | **Mod**  **SE** | **Emp**  **SE** | **Std**  **Bias** | **Bias** | **Mod**  **SE** | **Emp**  **SE** | **Std**  **Bias** |
| **Complete data (Cox)** | | -0.1 | 1.4 | 1.1 | -0.05 | -0.5 | 11.4 | 11.3 | -0.04 | -13.4 | 17.8 | 16.8 | -0.79 |
| **Complete data (Weibull)** | | 0.0 | 1.0 | 1.0 | 0.02 | -0.2 | 11.4 | 11.3 | -0.02 | 0.2 | 11.5 | 11.4 | 0.01 |
| **MCAR** | CCA | -0.2 | 2.7 | 1.8 | -0.09 | -1.4 | 16.3 | 16.6 | -0.08 | -26.9 | 29.2 | 28.7 | -0.93 |
|  | PMM | 0.4 | 2.1 | 1.5 | 0.27 | -0.5 | 15.4 | 13.5 | -0.03 | -19.4 | 21.9 | 22.2 | -0.87 |
|  | PMMSUBGP | -0.6 | 2.1 | 1.4 | -0.40 | 1.0 | 15.5 | 14.1 | 0.07 | -20.1 | 26.3 | 22.2 | -0.90 |
|  | PMMSUBGP* | -0.5 | 1.7 | 1.3 | -0.35 | 2.6 | 15.9 | 13.5 | 0.20 | -2.9 | 24.6 | 15.9 | -0.19 |
|  | PMMCOMP | 0.0 | 1.8 | 1.4 | -0.01 | -2.6 | 19.7 | 18.8 | -0.14 | -19.5 | 28.9 | 25.8 | -0.76 |
|  | LINMI | 4.1 | 4.5 | 2.3 | 1.81 | 8.6 | 19.8 | 14.1 | 0.61 | -30.3 | 53.3 | 26.4 | -1.15 |
| **MAR**  **(acute GvHD only)** | CCA | -0.7 | 2.4 | 1.4 | -0.51 | -13.6 | 11.9 | 12.0 | -1.13 | -6.7 | 22.7 | 21.4 | -0.31 |
|  | PMM | -0.2 | 1.6 | 1.4 | -0.13 | -0.1 | 11.5 | 11.3 | -0.01 | -13.6 | 17.8 | 16.8 | -0.81 |
|  | PMMSUBGP | -0.5 | 1.8 | 1.4 | -0.32 | 0.0 | 11.5 | 11.3 | 0.00 | -13.5 | 17.9 | 16.8 | -0.80 |
|  | PMMSUBGP* | -0.4 | 1.3 | 1.3 | -0.27 | 0.8 | 11.4 | 11.3 | 0.07 | -0.4 | 11.6 | 11.4 | -0.04 |
| **MNAR**  **(smallest acute GvHD only)** | CCA | 5.0 | 1.7 | 1.4 | 3.59 | -12.1 | 12.5 | 12.6 | -0.96 | -11.9 | 22.5 | 20.8 | -0.57 |
|  | PMM | 5.6 | 1.4 | 1.4 | 4.11 | -3.9 | 11.4 | 11.2 | -0.35 | -15.7 | 17.9 | 16.8 | -0.93 |
|  | PMMSUBGP | 5.5 | 1.7 | 1.5 | 3.76 | -3.8 | 11.4 | 11.2 | -0.34 | -15.6 | 17.9 | 16.8 | -0.93 |
|  | PMMSUBGP* | 5.8 | 1.2 | 1.4 | 4.18 | -3.8 | 11.4 | 11.1 | -0.34 | -2.0 | 11.6 | 11.4 | -0.17 |
| **MAR (relapse/ death only, 0→2 transition)** | CCA | 0.6 | 1.3 | 1.2 | 0.50 | 11.6 | 12.2 | 12.0 | 0.97 | -26.1 | 18.4 | 17.3 | -1.51 |
|  | PMM | 0.4 | 1.8 | 1.2 | 0.29 | -0.5 | 11.4 | 11.3 | -0.04 | -13.8 | 17.9 | 16.8 | -0.82 |
|  | PMMSUBGP | -0.1 | 1.4 | 1.2 | -0.05 | -0.5 | 11.4 | 11.3 | -0.05 | -13.3 | 17.8 | 16.8 | -0.79 |
|  | PMMSUBGP* | 0.0 | 1.0 | 1.1 | 0.04 | -0.2 | 11.4 | 11.3 | -0.02 | 0.2 | 11.5 | 11.4 | 0.02 |
| **MAR**  **(relapse/death only, both transitions)** | CCA | 0.0 | 2.5 | 1.4 | -0.01 | 10.7 | 16.9 | 16.4 | 0.65 | -28.4 | 24.1 | 22.4 | -1.27 |
|  | PMM | 0.1 | 1.9 | 1.3 | 0.06 | -1.2 | 19.6 | 16.9 | -0.07 | -16.6 | 25.1 | 22.8 | -0.73 |
|  | PMMSUBGP | -0.3 | 1.5 | 1.2 | -0.21 | 1.1 | 19.3 | 17.5 | 0.06 | -18.3 | 24.9 | 23.0 | -0.79 |
|  | PMMSUBGP* | -0.2 | 1.1 | 1.1 | -0.19 | 2.0 | 19.6 | 17.5 | 0.11 | -1.8 | 19.7 | 17.6 | -0.10 |
| **MNAR (smallest relapse/death only, both transitions)** | CCA | 4.1 | 1.4 | 1.3 | 3.19 | 38.9 | 13.7 | 14.0 | 2.77 | -56.9 | 19.0 | 18.3 | -3.11 |
|  | PMM | 12.9 | 7.6 | 4.4 | 2.92 | 5.5 | 11.9 | 12.1 | 0.45 | -32.3 | 19.1 | 17.7 | -1.82 |
|  | PMMSUBGP | 5.8 | 2.5 | 1.7 | 3.47 | 6.1 | 11.8 | 12.0 | 0.51 | -25.9 | 17.8 | 17.2 | -1.50 |
|  | PMMSUBGP* | 5.9 | 1.2 | 1.4 | 4.35 | 8.5 | 11.8 | 12.3 | 0.69 | -14.4 | 11.7 | 12.5 | -1.16 |
| **MAR (acute GvHD & relapse/death)** | CCA | -0.8 | 4.4 | 1.9 | -0.44 | -4.8 | 17.5 | 17.5 | -0.27 | -22.1 | 30.1 | 27.8 | -0.79 |
|  | PMM | 0.1 | 2.2 | 1.4 | 0.11 | -1.5 | 19.6 | 16.2 | -0.09 | -16.4 | 25.0 | 22.6 | -0.73 |
|  | PMMSUBGP | -0.5 | 2.2 | 1.5 | -0.36 | 2.6 | 20.1 | 16.3 | 0.16 | -20.1 | 25.5 | 22.5 | -0.89 |
|  | PMMSUBGP* | -0.4 | 1.4 | 1.3 | -0.35 | 1.9 | 19.7 | 15.9 | 0.12 | -1.5 | 19.8 | 16.0 | -0.09 |
| **MNAR (smallest acute GvHD) & MAR (relapse/death)** | CCA | 5.2 | 2.9 | 1.7 | 2.99 | -1.9 | 18.5 | 18.7 | -0.10 | -27.8 | 29.3 | 27.1 | -1.03 |
|  | PMM | 6.1 | 2.5 | 1.5 | 3.97 | -5.4 | 20.5 | 16.3 | -0.33 | -18.7 | 25.8 | 22.5 | -0.83 |
|  | PMMSUBGP | 5.6 | 1.7 | 1.5 | 3.74 | -1.9 | 20.0 | 16.1 | -0.12 | -21.5 | 25.4 | 22.6 | -0.95 |
|  | PMMSUBGP* | 5.8 | 1.3 | 1.4 | 4.16 | -2.1 | 20.4 | 16.6 | -0.13 | -3.7 | 20.5 | 16.8 | -0.22 |
| **MNAR (largest acute GvHD) & MAR (relapse/ death)** | CCA | -6.1 | 2.1 | 2.0 | -3.11 | -8.9 | 18.3 | 17.7 | -0.50 | -9.8 | 30.4 | 27.5 | -0.36 |
|  | PMM | -6.4 | 2.9 | 1.6 | -4.13 | 1.6 | 21.2 | 16.5 | 0.10 | -13.0 | 26.8 | 22.5 | -0.58 |
|  | PMMSUBGP | -6.3 | 2.0 | 1.7 | -3.62 | 5.0 | 20.1 | 16.0 | 0.31 | -16.7 | 25.6 | 21.9 | -0.76 |
|  | PMMSUBGP* | -8.3 | 0.9 | 0.9 | -9.07 | 9.3 | 19.9 | 16.2 | 0.57 | -1.0 | 20.0 | 16.3 | -0.06 |
| **MAR (acute GvHD) & MNAR (smallest relapse/ death)** | CCA | 4.5 | 2.6 | 1.6 | 2.82 | 33.2 | 15.1 | 15.9 | 2.08 | -57.4 | 24.1 | 22.4 | -2.57 |
|  | PMM | 13.4 | 7.7 | 4.7 | 2.88 | 5.9 | 11.9 | 12.0 | 0.49 | -33.2 | 19.1 | 17.8 | -1.87 |
|  | PMMSUBGP | 5.6 | 3.2 | 1.8 | 3.03 | 6.6 | 11.9 | 11.9 | 0.56 | -26.1 | 17.9 | 17.2 | -1.52 |
|  | PMMSUBGP* | 5.6 | 1.4 | 1.5 | 3.69 | 9.2 | 11.9 | 12.3 | 0.74 | -14.8 | 11.8 | 12.5 | -1.18 |
| **MAR (acute GvHD) & MNAR (largest relapse/ death)** | CCA | -2.9 | 1.4 | 1.3 | -2.23 | -104.9 | 3.6 | 5.5 | -19.04 | -443.5 | 10.9 | 12.6 | -35.14 |
|  | PMM | -0.6 | 1.4 | 1.3 | -0.49 | -95.1 | 4.7 | 6.1 | -15.48 | -454.9 | 5.1 | 11.8 | -38.39 |
|  | PMMSUBGP | -1.0 | 1.5 | 1.2 | -0.82 | -94.1 | 4.7 | 6.2 | -15.27 | -455.0 | 18.8 | 14.9 | -30.53 |
|  | PMMSUBGP* | -1.0 | 1.5 | 1.3 | -0.72 | -73.1 | 9.0 | 9.4 | -7.82 | 74.1 | 9.2 | 9.7 | 7.66 |
| **MNAR (smallest acute GvHD & smallest relapse/death)** | CCA | 11.6 | 1.9 | 1.6 | 7.19 | 30.0 | 15.7 | 16.2 | 1.85 | -60.5 | 23.9 | 22.5 | -2.69 |
|  | PMM | 19.4 | 7.5 | 4.5 | 4.31 | 0.8 | 11.8 | 11.9 | 0.07 | -34.1 | 19.0 | 17.5 | -1.94 |
|  | PMMSUBGP | 11.8 | 3.1 | 1.9 | 6.21 | 1.4 | 11.8 | 11.8 | 0.12 | -27.1 | 17.8 | 17.1 | -1.58 |
|  | PMMSUBGP* | 12.0 | 1.4 | 1.7 | 7.23 | 3.2 | 11.8 | 12.0 | 0.27 | -15.2 | 11.9 | 12.3 | -1.23 |

State indicators: 0 = transplanted; 1 = acute GvHD; 2 = relapse/death.

*Cox models were fit, except for methods indicated with a *, for which Weibull models were fit.

Monte Carlo SE for bias ranges from 0.03 to 0.14 for $e_{0}(2)$**,** from 0.17 to 0.59 for $e_{1}(2)$ and from 0.31 to 1.21 for $e_{2}(2)$.

PMM, MI by Type 1 predictive mean matching; PMMSUBGP, as for PMM with imputation models fit separately for patients with and without acute GvHD;

PMMCOMP, as for PMM, imputing time to acute GvHD and time from acute GvHD to relapse/death, with post-imputation calculation of time from transplant to relapse/death; LINMI, MI using draws from a linear imputation model, with imputation models fit separately for patients with and without acute GvHD.

### Section S4: Description of the National Health Service Cord Blood Bank (NHS CBB) dataset

Between 1996 and 2015, 432 first haematopoietic stem cell transplants from unrelated donors were performed using cord blood (CB) provided by the UK National Health Service (NHS) Cord Blood Bank (CBB). The patients included both adult and paediatric (aged 16 years or less) patients, treated for malignant and non-malignant blood disorders. The following (clinically relevant) baseline patient, donor, and transplant characteristics (with percentage missing data) were reported: number of CB units transplanted (0%) (single or double); patient age (0%); disease type (0%) (acute leukaemia, other blood cancer, non-malignant disorder) and disease status (35%) (in remission, relapse, other) at transplant; pre-transplant radiotherapy/chemotherapy regimen (4%) (intensive or reduced intensity); sex (1%) and cytomegalovirus-positive (CMV+) (12%) match between donor(s) and recipient; number of human leucocyte antigen (HLA) mismatches (13%) between donor(s) and recipient (well-matched: 0 or 1 mismatches; or not: 2 or more mismatches); dose at infusion (24%) (measured by the total nucleated cell count infused x 10^7^/kg patient weight, categorised as low, <3.0; medium, 3.0-5.0; high, >5.0 × 10^7^/kg). Year (0%) and country (0%) of transplant, and time of chronic GvHD (35%) were also reported.

Table S4a. Analysis of the motivating example. Hazard ratios (HR) and 95% confidence interval (CI) for covariates in the transition intensity model from transplant to acute GvHD.

| **Covariate (reference value)** |  | **Analysis approach** | | | | | | |
| --- | --- | --- | --- | --- | --- | --- | --- | --- |
|  | **CCA**  **(N=116)** | | **PMM**  **(N=432)** | | **PMMSUBGP**  **(N=432)** | | **PMMSUBGP Weibull**  **(N=432)** | |
|  | **HR** | **95% CI** | **HR** | **95% CI** | **HR** | **95% CI** | **HR** | **95% CI** |
| Double cord transplant (single) | 0.26 | 0.06-1.11 | 0.65 | 0.39-1.08 | 0.90 | 0.56-1.46 | 0.82 | 0.51-1.34 |
| Patient age (10-year increments) | 1.02 | 0.80-1.31 | 0.93 | 0.83-1.04 | 0.89 | 0.80-1.00 | 0.93 | 0.83-1.04 |
| Disease type (acute leukaemia) |  |  |  |  |  |  |  |  |
| Other blood cancer^1^ | 0.66 | 0.27-1.64 | 0.88 | 0.59-1.31 | 0.89 | 0.61-1.31 | 0.84 | 0.57-1.24 |
| Non-malignant disorder^2^ | 0.29 | 0.08-1.13 | 0.50 | 0.26-0.95 | 0.48 | 0.26-0.89 | 0.36 | 0.19-0.68 |
| Disease status at time of transplant  (in remission) |  |  |  |  |  |  |  |  |
| Relapse | 0.69 | 0.23-2.01 | 0.45 | 0.23-0.89 | 0.43 | 0.23-0.82 | 0.48 | 0.25-0.94 |
| Other^3^ | 1.50 | 0.49-4.56 | 1.20 | 0.70-2.06 | 1.25 | 0.75-2.07 | 1.27 | 0.75-2.15 |
| Reduced intensity conditioning regimen (intensive) | 1.86 | 0.88-3.93 | 1.38 | 0.97-1.96 | 1.36 | 0.96-1.93 | 1.39 | 0.98-1.99 |
| Donor-recipient CMV match (-/-) |  |  |  |  |  |  |  |  |
| -/+ | 1.40 | 0.65-3.00 | 1.36 | 0.87-2.11 | 1.27 | 0.82-1.97 | 1.45 | 0.92-2.30 |
| +/- | 1.45 | 0.64-3.29 | 1.46 | 0.95-2.27 | 1.29 | 0.83-2.01 | 1.49 | 0.93-2.39 |
| +/+ | 0.69 | 0.27-1.75 | 0.62 | 0.33-1.17 | 0.82 | 0.46-1.45 | 0.78 | 0.43-1.40 |
| Donor-recipient sex match (F/F) |  |  |  |  |  |  |  |  |
| F/M | 1.52 | 0.70-3.31 | 1.29 | 0.81-2.06 | 1.31 | 0.82-2.09 | 1.27 | 0.79-2.05 |
| M/F | 1.22 | 0.49-3.00 | 1.26 | 0.77-2.05 | 1.18 | 0.72-1.93 | 1.09 | 0.67-1.79 |
| M/M | 1.12 | 0.48-2.62 | 1.09 | 0.66-1.81 | 1.10 | 0.66-1.82 | 1.06 | 0.63-1.76 |
| Number of donor-recipient HLA mismatches^4^  (Well-matched: 0/1) |  |  |  |  |  |  |  |  |
| Not well-matched: 2 or more | 1.42 | 0.76-2.66 | 1.33 | 0.88-2.01 | 1.11 | 0.75-1.63 | 1.20 | 0.79-1.82 |
| TNC dose at infusion ×10^7^/kg (Low: <3.0) |  |  |  |  |  |  |  |  |
| Medium: 3.0-5.0 | 2.61 | 1.22-5.57 | 1.25 | 0.78-2.01 | 1.02 | 0.69-1.53 | 1.24 | 0.82-1.88 |
| High: > 5.0 | 1.46 | 0.61-3.47 | 1.15 | 0.74-1.79 | 0.84 | 0.54-1.32 | 1.01 | 0.64-1.58 |

Table S4b. Analysis of the motivating example. Hazard ratios (HR) and 95% confidence interval (CI) for covariates in the transition intensity model from transplant to relapse/death.

| **Covariate (reference value)** | **Analysis approach** | | | | | | | |
| --- | --- | --- | --- | --- | --- | --- | --- | --- |
|  | **CCA**  **(N=116)** | | **PMM**  **(N=432)** | | **PMMSUBGP (N=432)** | | **PMMSUBGP Weibull (N=432)** | |
|  | **HR** | **95% CI** | **HR** | **95% CI** | **HR** | **95% CI** | **HR** | **95% CI** |
| Double cord transplant (single) | 1.24 | 0.20-7.81 | 1.97 | 0.89-4.40 | 1.61 | 0.79-3.28 | 1.44 | 0.70-2.98 |
| Patient age (10-year increments) | 1.01 | 0.71-1.43 | 0.92 | 0.78-1.09 | 0.94 | 0.79-1.10 | 1.00 | 0.84-1.18 |
| Disease type (acute leukaemia) |  |  |  |  |  |  |  |  |
| Other blood cancer^1^ | 0.42 | 0.11-1.68 | 0.73 | 0.39-1.35 | 0.84 | 0.46-1.55 | 0.76 | 0.42-1.39 |
| Non-malignant disorder^2^ | 0.17 | 0.03-1.15 | 0.43 | 0.15-1.26 | 0.42 | 0.15-1.14 | 0.38 | 0.14-1.04 |
| Disease status at time of transplant  (in remission) |  |  |  |  |  |  |  |  |
| Relapse | 5.92 | 1.51-23.17 | 3.31 | 1.67-6.58 | 2.22 | 1.11-4.48 | 2.63 | 1.31-5.30 |
| Other^3^ | 3.17 | 0.61-16.59 | 1.86 | 0.77-4.49 | 1.72 | 0.77-3.84 | 1.73 | 0.77-3.88 |
| Reduced intensity conditioning regimen (intensive) | 0.67 | 0.23-2.00 | 0.89 | 0.52-1.51 | 0.93 | 0.56-1.54 | 0.94 | 0.57-1.57 |
| Donor-recipient CMV match (-/-) |  |  |  |  |  |  |  |  |
| -/+ | 1.30 | 0.42-4.02 | 1.32 | 0.58-3.03 | 1.60 | 0.74-3.44 | 1.71 | 0.78-3.73 |
| +/- | 0.81 | 0.25-2.68 | 0.77 | 0.32-1.85 | 1.15 | 0.51-2.57 | 1.34 | 0.59-3.04 |
| +/+ | 2.15 | 0.66-6.99 | 2.35 | 1.04-5.31 | 2.63 | 1.21-5.70 | 2.41 | 1.12-5.20 |
| Donor-recipient sex match (F/F) |  |  |  |  |  |  |  |  |
| F/M | 0.44 | 0.15-1.28 | 0.73 | 0.35-1.53 | 0.88 | 0.43-1.78 | 0.85 | 0.41-1.77 |
| M/F | 0.52 | 0.16-1.71 | 0.86 | 0.41-1.80 | 0.97 | 0.47-2.01 | 0.90 | 0.43-1.90 |
| M/M | 0.40 | 0.13-1.27 | 0.85 | 0.37-1.95 | 0.92 | 0.41-2.06 | 0.90 | 0.39-2.07 |
| Number of donor-recipient HLA mismatches^4^  (Well-matched: 0/1) |  |  |  |  |  |  |  |  |
| Not well-matched: 2 or more | 1.50 | 0.61-3.67 | 1.65 | 0.87-3.13 | 1.49 | 0.80-2.76 | 1.63 | 0.87-3.04 |
| TNC dose at infusion ×10^7^/kg  (Low: <3.0) |  |  |  |  |  |  |  |  |
| Medium: 3.0-5.0 | 1.88 | 0.63-5.63 | 1.14 | 0.56-2.31 | 1.06 | 0.55-2.06 | 1.18 | 0.61-2.31 |
| High: > 5.0 | 1.16 | 0.41-3.25 | 0.86 | 0.43-1.72 | 1.06 | 0.54-2.06 | 1.22 | 0.61-2.44 |

Table S4c. Analysis of the motivating example. Hazard ratios (HR) and 95% confidence interval (CI) for covariates in the transition intensity model from acute GvHD to relapse/death.

| **Covariate (reference value)** | **Analysis approach** | | | | | | | |
| --- | --- | --- | --- | --- | --- | --- | --- | --- |
|  | **CCA**  **(N=116)** | | **PMM**  **(N=432)** | | **PMMSUBGP (N=432)** | | **PMMSUBGP Weibull (N=432)** | |
|  | **HR** | **95% CI** | **HR** | **95% CI** | **HR** | **95% CI** | **HR** | **95% CI** |
| Double cord transplant (single) | 0.18 | 0.01-2.23 | 0.56 | 0.25-1.26 | 0.42 | 0.20-0.88 | 0.39 | 0.19-0.82 |
| Patient age (10-year increments) | 1.61 | 1.03-2.50 | 1.15 | 0.95-1.38 | 1.10 | 0.92-1.31 | 1.11 | 0.94-1.32 |
| Disease type (acute leukaemia) |  |  |  |  |  |  |  |  |
| Other blood cancer^1^ | 2.09 | 0.47-9.29 | 0.92 | 0.50-1.70 | 1.09 | 0.59-2.03 | 1.14 | 0.61-2.11 |
| Non-malignant disorder^2^ | 3.89 | 0.45-33.42 | 1.07 | 0.39-2.90 | 1.37 | 0.51-3.69 | 1.38 | 0.52-3.66 |
| Disease status at time of transplant  (in remission) |  |  |  |  |  |  |  |  |
| Relapse | 2.40 | 0.54-10.63 | 2.39 | 0.86-6.65 | 1.03 | 0.38-2.81 | 0.92 | 0.36-2.37 |
| Other^3^ | 1.29 | 0.19-8.64 | 1.63 | 0.70-3.75 | 1.14 | 0.50-2.57 | 1.03 | 0.47-2.27 |
| Reduced intensity conditioning regimen (intensive) | 0.40 | 0.09-1.85 | 0.78 | 0.44-1.41 | 0.99 | 0.54-1.83 | 0.99 | 0.54-1.81 |
| Donor-recipient CMV match (-/-) |  |  |  |  |  |  |  |  |
| -/+ | 0.86 | 0.29-2.57 | 1.68 | 0.81-3.47 | 1.90 | 0.94-3.82 | 1.93 | 0.97-3.85 |
| +/- | 1.11 | 0.31-3.92 | 1.33 | 0.62-2.86 | 1.29 | 0.62-2.71 | 1.27 | 0.61-2.62 |
| +/+ | 0.52 | 0.11-2.46 | 1.01 | 0.36-2.87 | 0.80 | 0.29-2.24 | 0.84 | 0.31-2.26 |
| Donor-recipient sex match (F/F) |  |  |  |  |  |  |  |  |
| F/M | 3.45 | 0.89-13.31 | 1.52 | 0.67-3.44 | 1.56 | 0.72-3.38 | 1.48 | 0.69-3.16 |
| M/F | 3.24 | 0.72-14.70 | 1.34 | 0.55-3.23 | 1.46 | 0.63-3.40 | 1.46 | 0.64-3.34 |
| M/M | 1.67 | 0.31-8.90 | 1.49 | 0.61-3.59 | 1.49 | 0.62-3.55 | 1.43 | 0.60-3.45 |
| Number of donor-recipient HLA mismatches^4^ (Well-matched: 0/1) |  |  |  |  |  |  |  |  |
| Not well-matched: 2 or more | 1.93 | 0.66-5.63 | 1.52 | 0.79-2.90 | 1.76 | 0.92-3.37 | 1.92 | 1.03-3.59 |
| TNC dose at infusion ×10^7^/kg  (Low: <3.0) |  |  |  |  |  |  |  |  |
| Medium: 3.0-5.0 | 0.82 | 0.24-2.74 | 0.80 | 0.41-1.58 | 0.62 | 0.36-1.08 | 0.65 | 0.38-1.11 |
| High: > 5.0 | 0.60 | 0.08-4.52 | 0.49 | 0.22-1.09 | 0.32 | 0.15-0.70 | 0.35 | 0.17-0.72 |

Table S4d. Analysis of the motivating example. Hazard ratios (HR) and 95% confidence interval (CI) for covariates in the transition intensity model from acute GvHD to relapse/death, including time from transplant to acute GvHD as a covariate.

| **Covariate (reference value)** | **Analysis approach** | | | | | | | |
| --- | --- | --- | --- | --- | --- | --- | --- | --- |
|  | **CCA**  **(N=116)** | | **PMM**  **(N=432)** | | **PMMSUBGP (N=432)** | | **PMMSUBGP Weibull (N=432)** | |
|  | **HR** | **95% CI** | **HR** | **95% CI** | **HR** | **95% CI** | **HR** | **95% CI** |
| Double cord transplant (single) | 0.21 | 0.02-2.55 | 0.56 | 0.24-1.26 | 0.42 | 0.20-0.88 | 0.39 | 0.19-0.83 |
| Patient age (10-year increments) | 1.52 | 0.96-2.39 | 1.15 | 0.95-1.38 | 1.10 | 0.92-1.31 | 1.11 | 0.93-1.32 |
| Disease type (acute leukaemia) |  |  |  |  |  |  |  |  |
| Other blood cancer^1^ | 1.58 | 0.36-7.00 | 0.93 | 0.50-1.73 | 1.09 | 0.59-2.03 | 1.13 | 0.61-2.09 |
| Non-malignant disorder^2^ | 2.32 | 0.25-21.65 | 1.08 | 0.39-3.00 | 1.37 | 0.51-3.72 | 1.35 | 0.51-3.59 |
| Disease status at time of transplant  (in remission) |  |  |  |  |  |  |  |  |
| Relapse | 2.40 | 0.53-10.96 | 2.39 | 0.86-6.66 | 1.03 | 0.38-2.82 | 0.92 | 0.36-2.31 |
| Other^3^ | 2.01 | 0.27-15.11 | 1.61 | 0.69-3.76 | 1.14 | 0.50-2.60 | 1.05 | 0.48-2.32 |
| Reduced intensity conditioning regimen (intensive) | 0.40 | 0.09-1.82 | 0.79 | 0.44-1.41 | 0.99 | 0.53-1.83 | 1.00 | 0.54-1.82 |
| Donor-recipient CMV match (-/-) |  |  |  |  |  |  |  |  |
| -/+ | 1.01 | 0.33-3.06 | 1.68 | 0.81-3.48 | 1.90 | 0.94-3.83 | 1.96 | 0.98-3.90 |
| +/- | 1.19 | 0.33-4.26 | 1.33 | 0.61-2.86 | 1.29 | 0.62-2.70 | 1.27 | 0.62-2.62 |
| +/+ | 0.60 | 0.12-2.92 | 1.01 | 0.36-2.86 | 0.80 | 0.29-2.25 | 0.85 | 0.32-2.30 |
| Donor-recipient sex match (F/F) |  |  |  |  |  |  |  |  |
| F/M | 4.81 | 1.11-20.76 | 1.52 | 0.67-3.44 | 1.56 | 0.71-3.43 | 1.52 | 0.71-3.26 |
| M/F | 4.53 | 0.92-22.16 | 1.33 | 0.55-3.22 | 1.46 | 0.63-3.42 | 1.49 | 0.65-3.39 |
| M/M | 1.90 | 0.35-10.26 | 1.47 | 0.61-3.58 | 1.49 | 0.62-3.58 | 1.44 | 0.60-3.48 |
| Number of donor-recipient HLA mismatches^4^ (Well-matched: 0/1) |  |  |  |  |  |  |  |  |
| Not well-matched: 2 or more | 1.63 | 0.56-4.76 | 1.52 | 0.79-2.91 | 1.76 | 0.92-3.38 | 1.90 | 1.02-3.54 |
| TNC dose at infusion ×10^7^/kg  (Low: <3.0) |  |  |  |  |  |  |  |  |
| Medium: 3.0-5.0 | 0.88 | 0.25-3.01 | 0.80 | 0.40-1.58 | 0.63 | 0.36-1.09 | 0.64 | 0.37-1.09 |
| High: > 5.0 | 0.54 | 0.07-4.02 | 0.49 | 0.22-1.09 | 0.32 | 0.15-0.70 | 0.34 | 0.16-0.72 |
| Time of acute GvHD | 1.01 | 0.99-1.03 | 1.00 | 0.99-1.01 | 1.00 | 0.99-1.01 | 1.00 | 0.99-1.01 |

CMV, cytomegalovirus; HLA, human leucocyte antigen; TNC, total nucleated cells. (Footnotes continued overleaf)

Unless otherwise stated, Cox transition intensity models were fitted.

CCA, complete case analysis; PMM, MI by Type 1 predictive mean matching;

PMMSUBGP, as for PMM with imputation models fit separately for patients experiencing (i) both acute and chronic GvHD or chronic GvHD without acute GvHD, (ii) acute GvHD without chronic GvHD, (iii) relapse without GvHD or neither GvHD nor relapse.

^1^ Other blood cancer includes lymphoproliferative and plasma cell disorders, myelodysplastic syndromes and myeloproliferative disorders.

^2^ Non-malignant disorder includes histiocytic disorder, solid tumour, bone marrow failure syndrome, haemoglobinopathy, primary immune deficiency and inborn error of metabolism.

^3^ Other disease status includes acute, chronic and accelerated phase, refractory disease, transformed to acute leukaemia, blastic crisis, myelodysplastic syndromes, myeloproliferative disorders, and non-malignant disorders.

^4^ HLA A and B loci at antigenic level and DR-B1 at allelic level.

**Section S5. R code to generate data for the simulation study**

1. **Generating data for a three-state Markov MSM**

##First competing risks experiment

#hazard of aGvHD

k_agvhd=1.5

l_agvhd=36

#hazard of relapse/death

k_rel_death=0.9

l_rel_death=120

#also add two covariates

#1. in relapse or not

#2. number of cords received

relapse=rbinom(500000,1,0.2)

doublecord=rbinom(500000,1,0.45)

\# add sampno

sampno=rep(1:1000, 500)

covs_all = data.frame(id = 1:500000, relapse, doublecord, sampno)

#simulate fup to 5 years for all pts (censor after all times drawn)

#1. not in relapse

h_all = function(t,x,betas)#function needs this structure in simsurv

((k_agvhd/l_agvhd)*(t/l_agvhd)^(k_agvhd-1)

+

(k_rel_death/l_rel_death)*(t/l_rel_death)^(k_rel_death-1))

covs=data.frame(id = covs_all[covs_all$relapse==0,1])

simdata = simsurv(hazard = h_all,maxt=365,x=covs,seed=4752)

#assume order of rows is unchanged and drop id from this dataset and merge back in id from covs

simdata_1=cbind(simdata[,2:3],covs_all[covs_all$relapse==0,])

#now calculate prob of agvhd for binomial draws to determine whether event of interest or #competing event

simdata_1$h_all=h_all(t=simdata_1$eventtime)

h_agvhd = function(t)

((k_agvhd/l_agvhd)*(t/l_agvhd)^(k_agvhd-1))

simdata_1$h_agvhd=h_agvhd(t=simdata_1$eventtime)

#now calculate prob of agvhd at event time

simdata_1$p_agvhd=simdata_1$h_agvhd/simdata_1$h_all

#2. in relapse

#use simsurv

h_all2 = function(t,x,betas)#function needs this structure in simsurv

(exp(-0.8)*(k_agvhd/l_agvhd)*(t/l_agvhd)^(k_agvhd-1)

+

exp(1.2)*(k_rel_death/l_rel_death)*(t/l_rel_death)^(k_rel_death-1))

covs2=data.frame(id = covs_all[covs_all$relapse==1,1])

simdata2 = simsurv(hazard = h_all2,maxt=365,x=covs2,seed=78947)

#assume order of rows is unchanged and drop id from this dataset and merge back in id from covs

simdata2_1=cbind(simdata2[,2:3],covs_all[covs_all$relapse==1,])

#now calculate prob of agvhd for binomial draws to determine whether event of interest or #competing event

simdata2_1$h_all=h_all2(t=simdata2_1$eventtime)

h_agvhd2 = function(t)

(exp(-0.8)*(k_agvhd/l_agvhd)*(t/l_agvhd)^(k_agvhd-1))

simdata2_1$h_agvhd=h_agvhd2(t=simdata2_1$eventtime)

#now calculate prob of agvhd at event time

simdata2_1$p_agvhd=simdata2_1$h_agvhd/simdata2_1$h_all

\##### Combine all rows #####

simdata_all=rbind(simdata_1,simdata2_1)

###Now run binomial experiment to determine event type

for (i in 1:500000) {

simdata_all$agvhd_status[i]=ifelse(rbinom(1,1,simdata_all$p_agvhd[i])==1,1,0)

}

#rename eventtime for consistency with other progs

names(simdata_all)[names(simdata_all) == 'eventtime'] = 'agvhd_time'

#Need to create relapse time and status cols

simdata_all$relapse_or_death_time=simdata_all$agvhd_time

simdata_all$relapse_or_death_status=ifelse(simdata_all$agvhd_status==0,1,0)

#order by id

simdata_all=simdata_all[order(simdata_all$id),]

####Now calculate event times for relapse/death after aGvHD

#Create transition 1->2 times

#Based on conditional survival model

###For simplicity, calculate for all cases, but discard rows for cases with agvhd_status=0

#Weibull params

k_rel_death12=0.8

l_rel_death12=160

#First calculate S(t0) ie at time of acute GvHD

simdata_all$S_t0=exp(-((simdata_all$agvhd_time/l_rel_death12)^k_rel_death12)*exp(1.2*simdata_all$relapse - simdata_all$doublecord))

#Generate times using inverse transform sampling and

#discard relapse times with agvhd_status=0 i.e. already in relapse/death state

simdata_all$runif=runif(500000)

simdata_all$rel_death_time=ifelse(simdata_all$agvhd_status==1,

160*((-log(simdata_all$runif*simdata_all$S_t0)/

exp(1.2*simdata_all$relapse - simdata_all$doublecord))^1.25),

simdata_all$relapse_or_death_time)

#Set all rel_death_status to 1 as no censored times at this point

simdata_all$rel_death_status=1

###Now generate censoring times by sampling from uniform dist

#Since all transitions from transplant occurred within the year, only 1-2 transitions will be #censored

simdata_all$cens_time=runif(500000, min = 365, max = 1826)

#update relapse/death time and status for these cases

simdata_all$rel_death_time_uncens=simdata_all$rel_death_time

simdata_all$rel_death_time=ifelse(simdata_all$rel_death_time_uncens>simdata_all$cens_time, simdata_all$cens_time, simdata_all$rel_death_time_uncens)

simdata_all$rel_death_status_uncens=simdata_all$rel_death_status

simdata_all$rel_death_status=ifelse(simdata_all$rel_death_time_uncens>simdata_all$cens_time, 0,simdata_all$rel_death_status_uncens)

2. **Missing data mechanisms and MI methods**

**#Functions for Rubin's rules to obtain SE (two versions, for parameter ests and RELOS)**

rubins_SE=function(n) {

SE_est=sqrt(mean(imp_results[,n+4]^2,na.rm=T)+((1+1/nimp)*1/(nimp-1)*sum((imp_results[,n]-mean(imp_results[,n],na.rm=T))^2,na.rm=T)))

return(SE_est)

}

rubins_SE2=function(n) {

SE_est=sqrt(mean(imp_results[,n+3]^2,na.rm=T)+((1+1/nimp)*1/(nimp-1)*sum((imp_results[,n]-mean(imp_results[,n],na.rm=T))^2,na.rm=T)))

return(SE_est)

}

**#Missing data mechanisms**

for (i in 1:1000)

{

sample1=simdata_all[simdata_all$sampno==i,]

#MCAR

\# For each event, set 30% of times to missing

for (j in 1:500){

sample1$rel_death_time_MCAR30[j]=ifelse(rbinom(1,1,0.3)==1,NA,sample1$rel_death_time[j])

sample1$agvhd_time_MCAR30[j]=ifelse(rbinom(1,1,0.3)==1,NA,sample1$agvhd_time[j])

}

#MAR – to avoid repetition, only one example of a MAR mechanism is shown

sample1$agvhd_time_MAR30=sample1$agvhd_time

for (j in 1:500){

if (sample1$agvhd_status[j]==1 & rbinom(1,1,0.2*(1 + sample1$doublecord[j]))==1)

{ sample1$agvhd_time_MAR30[j]=NA}

}

#MNAR – to avoid repetition, only one example of a MNAR mechanism is shown

sample1=sample1[order(-sample1$rel_death_status,sample1$rel_death_time),]

sample1$rel_death_time_MNAR30=sample1$rel_death_time

sample1$rel_death_time_MNAR30[1:round(0.3*nrow(sample1[sample1$rel_death_status==1,]))]=NA

for (j in 1:500){

\# to set up correctly for use with mstate, also need to set the associated agvhd_time to missing

\## for pts who go from tx->rel/death

sample1$agvhd_time_MNAR30[j]=ifelse(sample1$agvhd_status[j]==0 & is.na(sample1$rel_death_time_MNAR30[j])==T,

NA,sample1$agvhd_time[j])

}

**# MI methods – delete as appropriate**

midata=sample1[,c(2:3,6,8,12:13)]

#PMM

imp=mice(midata,m=nimp,print=FALSE)

#PMMSUBGP

#agvhd_status=0

predMatrix_0<-matrix(rep(0,25),ncol=5)

colnames(predMatrix_0)<-rownames(predMatrix_0)<-names(midata[,1:5])

predMatrix_0["rel_death_time_MCAR30",] <-c(1,1,0,0,0)

imp_0=mice(midata[midata$agvhd_status==0,1:5],m=nimp,predictorMatrix=predMatrix_0, print=FALSE)

#agvhd_status=1

predMatrix_1<-matrix(rep(0,36),ncol=6)

colnames(predMatrix_1)<-rownames(predMatrix_1)<-names(midata)

predMatrix_1["agvhd_time_MCAR30",] <-c(1,1,0,1,1,0)

predMatrix_1["rel_death_time_MCAR30",] <-c(1,1,0,1,0,1)

imp_1=mice(midata[midata$agvhd_status==1,],m=nimp,predictorMatrix=predMatrix_1, print=FALSE)

#PMMCOMP

#Add in time from agvhd to relapse for imputation purposes

sample1$agvhd_to_relapse_timeMCAR30=sample1$rel_death_time_MCAR30-sample1$agvhd_time_MCAR30

midata=sample1[,c(2:3,6,8,13:14)]

#agvhd_status=0

predMatrix_0<-matrix(rep(0,25),ncol=5)

colnames(predMatrix_0)<-rownames(predMatrix_0)<-names(midata[,1:5])

predMatrix_0["agvhd_time_MCAR30",] <-c(1,1,0,0,0)

#exclude agvhd_to_relapse_timeMCAR30 for agvhd_status=0 as n/a

imp_0=mice(midata[midata$agvhd_status==0,1:5],m=nimp,predictorMatrix=predMatrix_0, print=FALSE)

#agvhd_status=1

predMatrix_1<-matrix(rep(0,36),ncol=6)

colnames(predMatrix_1)<-rownames(predMatrix_1)<-names(midata)

predMatrix_1["agvhd_time_MCAR30",] <-c(1,1,0,1,0,1)

predMatrix_1["agvhd_to_relapse_timeMCAR30",] <-c(1,1,0,1,1,0)

imp_1=mice(midata[midata$agvhd_status==1,],m=nimp,predictorMatrix=predMatrix_1, print=FALSE)

#LINMI

#agvhd_status=0

predMatrix_0<-matrix(rep(0,36),ncol=6)

colnames(predMatrix_0)<-rownames(predMatrix_0)<-names(midata)

predMatrix_0["rel_death_time_MCAR30",] <-c(1,1,0,0,0,0)

imp_0=mice(midata[midata$agvhd_status==0,],m=nimp,method=c('','','','','norm',''),

predictorMatrix=predMatrix_0, print=FALSE)

#agvhd_status=1

predMatrix_1<-matrix(rep(0,36),ncol=6)

colnames(predMatrix_1)<-rownames(predMatrix_1)<-names(midata)

predMatrix_1["rel_death_time_MCAR30",] <-c(1,1,0,1,0,1)

predMatrix_1["agvhd_time_MCAR30",] <-c(1,1,0,1,1,0)

imp_1=mice(midata[midata$agvhd_status==1,],m=nimp,method=c('','','','','norm','norm'),

predictorMatrix=predMatrix_1, print=FALSE)

\# Run desired analysis for each imputed dataset

imp_results = data.frame(relapse.1=0,relapse.2=0,relapse.3=0,doublecord.3=0,

relapse.1SE=0,relapse.2SE=0,relapse.3SE=0,doublecord.3SE=0,agvhd_time=0, agvhd_timeSE=0,ELOS1=0,ELOS2=0,ELOS3=0,ELOS_SE1=0,ELOS_SE2=0,ELOS_SE3=0)

for (m in 1:nimp)

{

#recombine imputed data as required before analysis e.g. for PMMCOMP:

complete_0=complete(imp_0,m)

complete_0$rel_death_time_MCAR30=complete_0$agvhd_time_MCAR30

complete_1=complete(imp_1,m)

complete_1$rel_death_time_MCAR30=complete_1$agvhd_to_relapse_timeMCAR30 + complete_1$agvhd_time_MCAR30

complete=rbind.data.frame(complete_0,complete_1[,-6])

##Run required analysis model – see next section##

}

#Apply Rubin's rules

results[i,c(1:4,10:12)]=apply(imp_results[,c(1:4,11:13)],2,mean,na.rm=T)

#Rubins rules for gamma12

mean_gam12=mean(imp_results[,9],na.rm=T)

SE_gam12=sqrt(mean(imp_results[,10]^2)+((1+1/nimp)*1/(nimp-1)*sum((imp_results[,9]-mean(imp_results[,9],na.rm=T))^2)))

results[i,9]=ifelse(mean_gam12 - 1.96*SE_gam12<0 & mean_gam12 + 1.96*SE_gam12>0,1,0)

results[i,5:8]=lapply(1:4,rubins_SE)

results[i,13:15]=lapply(11:13,rubins_SE2)

}

3. **MSM analysis**

**#1. Cox model for each transition intensity**

**#1.1. Function for RELOS SE**

RELOS_SE_boot <- function(data,nboot,agvhdtime,reldeathtime) {

library(survival)

library(mstate)

**#Adapting ‘intccr’ bssmle_se R code**

\# **Park J, Bakoyannis G, Yiannoutsos C. Semiparametric competing risks regression under # interval censoring using the R package intccr. *Comput Methods Programs Biomed*.**

**# 2019;173:167-176**

tmp <- data.frame()

for(k in 1:nboot){

samp=data[sample(dim(data)[1], replace = TRUE),]

samp$boot=k

tmp <- rbind(tmp,samp)

}

m <- NULL

tmat <- transMat(x = list(c(2, 3), c(3), c()), names = c("Tx", "aGvHD", "Rel/Death"))

res.bt=data.frame(ELOS1=0,ELOS2=0,ELOS3=0)

for(m in 1:nboot){

#prep dset in 'long' format

tmplong <- mstate::msprep(data = tmp[tmp$boot==m,], trans = tmat,

time = c(NA, agvhdtime, reldeathtime),

status = c(NA, "agvhd_status", "rel_death_status"),

keep = c("doublecord","relapse"))

covs <- c("doublecord", "relapse")

tmplong_cov <- mstate::expand.covs(tmplong, covs, longnames = FALSE)

#allow for errors

skip_to_next <- FALSE

tryCatch(survival::coxph(Surv(Tstart, Tstop, status) ~ relapse.1 + relapse.2 + relapse.3 + doublecord.3 + strata(trans), data = tmplong_cov, method = "breslow"),

error = function(e) { skip_to_next <<- TRUE},

warning = function(w) {skip_to_next <<- TRUE})

if(skip_to_next) { next }

tmpfit=NULL

tmpfit=survival::coxph(Surv(Tstart, Tstop, status) ~ relapse.1 + relapse.2 + relapse.3 + doublecord.3 + strata(trans), data = tmplong_cov, method = "breslow")

baseline <- data.frame(trans=1:3,relapse.1=c(0,0,0),relapse.2=c(0,0,0),relapse.3=c(0,0,0),

doublecord.3=c(0,0,0),strata=1:3)

tmpmsf = mstate::msfit(tmpfit, baseline, trans = tmat)

#calc probs

tmppt=mstate::probtrans(tmpmsf, predt = 0, variance=FALSE)

tmpELOS=data.frame(ELOS1=0,ELOS2=0,ELOS3=0)

for (j in 1:(length(tmppt[[1]]$time[tmppt[[1]]$time<731])-1))

{

tmpELOS$ELOS1=tmpELOS$ELOS1+(tmppt[[1]]$pstate1[j]*(tmppt[[1]]$time[j+1]-tmppt[[1]]$time[j]))

tmpELOS$ELOS2=tmpELOS$ELOS2+(tmppt[[1]]$pstate2[j]*(tmppt[[1]]$time[j+1]-tmppt[[1]]$time[j]))

tmpELOS$ELOS3=tmpELOS$ELOS3+(tmppt[[1]]$pstate3[j]*(tmppt[[1]]$time[j+1]-tmppt[[1]]$time[j]))

}

res.bt[m,]=tmpELOS

}

result <- sqrt(diag(var(res.bt[,1:3],na.rm=T)))

return(result)

}

**##1.2 Complete case analysis – for MI, first impute data then proceed as for CCA ##**

results = data.frame(relapse.1=0,relapse.2=0,relapse.3=0,doublecord.3=0, relapse.1SE=0,relapse.2SE=0,relapse.3SE=0,doublecord.3SE=0,

agvhd_time_coverage=0,

ELOS1=0,ELOS2=0,ELOS3=0,ELOS_SE1=0,ELOS_SE2=0,ELOS_SE3=0)

for (i in 1:1000)

{

sample1=simdata_all[simdata_all$sampno==i,]

tmat <- transMat(x = list(c(2, 3), c(3), c()), names = c("Tx", "aGvHD", "Rel/Death"))

sample1_long <- msprep(data = sample1, trans = tmat, time = c(NA, "agvhd_time", "rel_death_time"),

status = c(NA, "agvhd_status", "rel_death_status"),

keep = c("doublecord","relapse","sampno","agvhd_time"), id="id")

covs <- c("doublecord", "relapse")

sample1_long_cov <- expand.covs(sample1_long, covs, longnames = FALSE)

fit <- coxph(Surv(Tstart, Tstop, status) ~ relapse.1 + relapse.2 + relapse.3 + doublecord.3 + strata(trans), data = sample1_long_cov, method = "breslow")

results[i,1:4]=summary(fit)$coefficients[1:4]

results[i,5:8]=sqrt(diag(vcov(fit)))

#gamma12

fit12 <- coxph(Surv(Tstart, Tstop, status) ~ relapse + doublecord +	agvhd_time,

data = sample1_long, method = "breslow", subset=(trans=="3"))

results[i,9]=ifelse(summary(fit12)$coefficients[3,5]>=0.05,1,0)

#RELOS

baseline <-	data.frame(trans=1:3,relapse.1=c(0,0,0),relapse.2=c(0,0,0),relapse.3=c(0,0,0),

doublecord.3=c(0,0,0),strata=1:3)

msf0 <- msfit(fit, baseline, trans = tmat)

#calc trans probs

pt0=probtrans(msf0, predt = 0, variance=FALSE)

ELOS=data.frame(ELOS1=0,ELOS2=0,ELOS3=0)

for (j in 1:(length(pt0[[1]]$time[pt0[[1]]$time<731])-1))

{

ELOS$ELOS1=ELOS$ELOS1+(pt0[[1]]$pstate1[j]*(pt0[[1]]$time[j+1]-pt0[[1]]$time[j]))

ELOS$ELOS2=ELOS$ELOS2+(pt0[[1]]$pstate2[j]*(pt0[[1]]$time[j+1]-pt0[[1]]$time[j]))

ELOS$ELOS3=ELOS$ELOS3+(pt0[[1]]$pstate3[j]*(pt0[[1]]$time[j+1]-pt0[[1]]$time[j]))

}

results[i,10:12]=ELOS

results[i,13:15]=RELOS_SE_boot(data=sample1,nboot=50,"agvhd_time", "rel_death_time")}

**#2. Weibull model for each transition intensity**

**#2.1. Define function for ELOS SE**

ELOS_delta=function(obj){

msf0 <- msfit.flexsurvreg(obj, t=seq(0,730,by=0.1), trans=tmat, newdata=baseline,variance=FALSE)

#calc trans probs

pt0=probtrans(msf0, predt = 0, variance=FALSE)

ELOS=data.frame(ELOS1=0,ELOS2=0,ELOS3=0)

for (j in 1:(length(pt0[[1]]$time[pt0[[1]]$time<731])-1))

{

ELOS$ELOS1=ELOS$ELOS1+(pt0[[1]]$pstate1[j]*(pt0[[1]]$time[j+1]-pt0[[1]]$time[j]))

ELOS$ELOS2=ELOS$ELOS2+(pt0[[1]]$pstate2[j]*(pt0[[1]]$time[j+1]-pt0[[1]]$time[j]))

ELOS$ELOS3=ELOS$ELOS3+(pt0[[1]]$pstate3[j]*(pt0[[1]]$time[j+1]-pt0[[1]]$time[j]))

}

return(ELOS)

}

**##2.2 Complete data analysis – for MI, first impute data then proceed as for CCA ##**

results = data.frame(relapse.1=0,relapse.2=0,relapse.3=0,doublecord.3=0,

relapse.1SE=0,relapse.2SE=0,relapse.3SE=0,doublecord.3SE=0,

agvhd_time_coverage=0,

ELOS1=0,ELOS2=0,ELOS3=0,ELOS_SE1=0,ELOS_SE2=0,ELOS_SE3=0)

for (i in 1:1000)

{

sample1=simdata_all [simdata_all$sampno==i,]

tmat <- transMat(x = list(c(2, 3), c(3), c()), names = c("Tx", "aGvHD", "Rel/Death"))

sample1_long <- msprep(data = sample1, trans = tmat, time = c(NA, "agvhd_time", "rel_death_time"),

status = c(NA, "agvhd_status", "rel_death_status"),

keep = c("doublecord","relapse","sampno","agvhd_time"), id="id")

fit.list <- vector(3, mode="list")

fit.list[[1]]=flexsurvreg(Surv(Tstart, Tstop, status) ~ relapse, subset = (trans == 1), data = sample1_long,dist ="weibullPH",

inits=c(shape=1.5,scale=36^(-1.5)))

fit.list[[2]]=flexsurvreg(Surv(Tstart, Tstop, status) ~ relapse, subset = (trans == 2), data = sample1_long,dist = "weibullPH", inits=c(shape=0.9,scale=120^(-0.9)))

fit.list[[3]]=flexsurvreg(Surv(Tstart, Tstop, status) ~ relapse + doublecord, subset = (trans == 3),

data = sample1_long,dist = "weibullPH", inits=c(shape=0.8,scale=160^(-0.8)))

results[i,1]=fit.list[[1]]$res[3,1]

results[i,2]=fit.list[[2]]$res[3,1]

results[i,3:4]=fit.list[[3]]$res[3:4,1]

#SE

results[i,5]=fit.list[[1]]$res[3,4]

results[i,6]=fit.list[[2]]$res[3,4]

results[i,7:8]=fit.list[[3]]$res[3:4,4]

#gamma12

fit12=flexsurvreg(Surv(Tstart, Tstop, status) ~ relapse + doublecord + agvhd_time, subset = (trans == 3), data = sample1_long, dist = "weibullPH", inits=c(shape=0.8,scale=160^(-0.8)))

results[i,9]=ifelse(fit12$res[5,2]<0 & fit12$res[5,3]>0,1,0)

#now apply msfit to obtain estimates of ELOS

#Define pt with baseline values of covariates

baseline <- data.frame(trans=1:3,relapse=c(0,0,0),doublecord=c(0,0,0))

msf0 <- msfit.flexsurvreg(fit.list, t=seq(0,730,by=0.1), trans=tmat, newdata=baseline,variance=FALSE)

#calc trans probs

pt0=probtrans(msf0, predt = 0, variance=FALSE)

ELOS=data.frame(ELOS1=0,ELOS2=0,ELOS3=0)

for (j in 1:(length(pt0[[1]]$time[pt0[[1]]$time<731])-1))

{

ELOS$ELOS1=ELOS$ELOS1+(pt0[[1]]$pstate1[j]*(pt0[[1]]$time[j+1]-pt0[[1]]$time[j]))

ELOS$ELOS2=ELOS$ELOS2+(pt0[[1]]$pstate2[j]*(pt0[[1]]$time[j+1]-pt0[[1]]$time[j]))

ELOS$ELOS3=ELOS$ELOS3+(pt0[[1]]$pstate3[j]*(pt0[[1]]$time[j+1]-pt0[[1]]$time[j]))

}

results[i,10:12]=ELOS[1:3]

#SE using the delta method

#Obtaining sigma

#note using res not coef as want to keep params on original scale

coef=c(as.numeric(fit.list[[1]]$res[1:2,1]),as.numeric(fit.list[[2]]$res[1:2,1]),

as.numeric(fit.list[[3]]$res[1:2,1]))

#need to transform each weibull param est from cov as only shown in log- #transformed mode

#using delta method

cov=matrix(rep(0,36),ncol=6)#assuming independence between strata

cov[1,1]=(coef[1])^2*(as.numeric(fit.list[[1]]$cov[1,1]))

cov[1,2]=coef[1]*coef[2]*(as.numeric(fit.list[[1]]$cov[1,2]))

cov[2,1]=cov[1,2]

cov[2,2]=(coef[2])^2*(as.numeric(fit.list[[1]]$cov[2,2]))

cov[3,3]=(coef[3])^2*(as.numeric(fit.list[[2]]$cov[1,1]))

cov[3,4]=coef[3]*coef[4]*(as.numeric(fit.list[[2]]$cov[1,2]))

cov[4,3]=cov[3,4]

cov[4,4]=(coef[4])^2*(as.numeric(fit.list[[2]]$cov[2,2]))

cov[5,5]=(coef[5])^2*(as.numeric(fit.list[[3]]$cov[1,1]))

cov[5,6]=coef[5]*coef[6]*(as.numeric(fit.list[[3]]$cov[1,2]))

cov[6,5]=cov[5,6]

cov[6,6]=(coef[6])^2*(as.numeric(fit.list[[3]]$cov[2,2]))

#Calculate partial derivatives using finite differences

eps=0.00001

#Initialise dataset

partial=matrix(rep(0,18),ncol=3)

fit.listu1=fit.list

#fit.listu1[[1]]$res.t[1,1]=fit.list[[1]]$res.t[1,1]+eps

fit.listu1[[1]]$res.t[1,1]=log(fit.list[[1]]$res[1,1]+eps)

#change param by eps on original scale

fit.listl1=fit.list

#fit.listl1[[1]]$res.t[1,1]=fit.list[[1]]$res.t[1,1]-eps

fit.listl1[[1]]$res.t[1,1]=log(fit.list[[1]]$res[1,1]-eps)

partial[1,1:3]=c(as.numeric((ELOS_delta(fit.listu1)-ELOS_delta(fit.listl1))/(2*eps)))[1:3]

fit.listu2=fit.list

#fit.listu2[[1]]$res.t[2,1]=fit.list[[1]]$res.t[2,1]+eps

fit.listu2[[1]]$res.t[2,1]=log(fit.list[[1]]$res[2,1]+eps)

fit.listl2=fit.list

#fit.listl2[[1]]$res.t[2,1]=fit.list[[1]]$res.t[2,1]-eps

fit.listl2[[1]]$res.t[2,1]=log(fit.list[[1]]$res[2,1]-eps)

partial[2,1:3]=c(as.numeric((ELOS_delta(fit.listu2)-ELOS_delta(fit.listl2))/(2*eps)))[1:3]

fit.listu3=fit.list

#fit.listu3[[2]]$res.t[1,1]=fit.list[[2]]$res.t[1,1]+eps

fit.listu3[[2]]$res.t[1,1]=log(fit.list[[2]]$res[1,1]+eps)

fit.listl3=fit.list

#fit.listl3[[2]]$res.t[1,1]=fit.list[[2]]$res.t[1,1]-eps

fit.listl3[[2]]$res.t[1,1]=log(fit.list[[2]]$res[1,1]-eps)

partial[3,1:3]=c(as.numeric((ELOS_delta(fit.listu3)-ELOS_delta(fit.listl3))/(2*eps)))[1:3]

fit.listu4=fit.list

#fit.listu4[[2]]$res.t[2,1]=fit.list[[2]]$res.t[2,1]+eps

fit.listu4[[2]]$res.t[2,1]=log(fit.list[[2]]$res[2,1]+eps)

fit.listl4=fit.list

#fit.listl4[[2]]$res.t[2,1]=fit.list[[2]]$res.t[2,1]-eps

fit.listl4[[2]]$res.t[2,1]=log(fit.list[[2]]$res[2,1]-eps)

partial[4,1:3]=c(as.numeric((ELOS_delta(fit.listu4)-ELOS_delta(fit.listl4))/(2*eps)))[1:3]

fit.listu5=fit.list

#fit.listu5[[3]]$res.t[1,1]=fit.list[[3]]$res.t[1,1]+eps

fit.listu5[[3]]$res.t[1,1]=log(fit.list[[3]]$res[1,1]+eps)

fit.listl5=fit.list

#fit.listl5[[3]]$res.t[1,1]=fit.list[[3]]$res.t[1,1]-eps

fit.listl5[[3]]$res.t[1,1]=log(fit.list[[3]]$res[1,1]-eps)

partial[5,1:3]=c(as.numeric((ELOS_delta(fit.listu5)-ELOS_delta(fit.listl5))/(2*eps)))[1:3]

fit.listu6=fit.list

#fit.listu6[[3]]$res.t[2,1]=fit.list[[3]]$res.t[2,1]+eps

fit.listu6[[3]]$res.t[2,1]=log(fit.list[[3]]$res[2,1]+eps)

fit.listl6=fit.list

#fit.listl6[[3]]$res.t[2,1]=fit.list[[3]]$res.t[2,1]-eps

fit.listl6[[3]]$res.t[2,1]=log(fit.list[[3]]$res[2,1]-eps)

partial[6,1:3]=c(as.numeric((ELOS_delta(fit.listu6)-ELOS_delta(fit.listl6))/(2*eps)))[1:3]

results[i,13]=sqrt(t(partial[,1]) %*% cov %*% partial[,1])

results[i,14]=sqrt(t(partial[,2]) %*% cov %*% partial[,2])

results[i,15]=sqrt(t(partial[,3]) %*% cov %*% partial[,3])

#results[i,5:10]=ELOS

}
